# Signatures of the neutralizing dengue virus-reactive memory B cell repertoire identified by multiplexed envelope dimer probes

**DOI:** 10.64898/2026.09.15.26363039

**Authors:** Kelsey E. Lowman, Rosemary A. Aogo, Sarah Smith, Ysabelle Broderson, Saba Firdous, Gitanjali Bhushan, Devina J. Thiono, Patrick Mpingabo, Amy R. Henry, Timothy S. Johnston, Jesmine Roberts-Torres, Luis Estrada, Chaim A. Schramm, Foo Cheung, Jessie J. Polanco, Richard Apps, Krista Gangler, Jeffrey I. Cohen, Sonya Jacobson, Edgar Davidson, Benjamin J. Doranz, James Cherry, Iyadh Douagi, Barbara A. Blacklaws, Jonathan L. Heeney, Aravinda M. de Silva, Daniel C. Douek, Leah C. Katzelnick

## Abstract

Sequential dengue virus (DENV) infection is thought to induce long-lived protection against dengue. However, little is known about the specificity, phenotype, and repertoire of the enduring B cell response due to limitations in high-throughput methods. We used stabilized DENV envelope (E) dimers as probes for in-depth memory B cell repertoire characterization using two complementary sequencing strategies. Using Rapid Assembly, Transfection, and Production of Immunoglobulins (RATP-Ig), we isolated 227 DENV2 E dimer-specific memory B cells from an immune donor, of which 87% bound at least two serotypes, 68% bound the fusion loop (FL-sensitive), and 42% were neutralizing. Among the most neutralizing, cross-binding antibodies, the FL-sensitive antibodies had weaker but broad neutralization, including to other flaviviruses, whereas FL-‘agnostic’ antibodies mapped to E domain II and III and were potently neutralizing, but only against one or two serotypes. Competitive binding of the DENV1-4 E dimers distinguished the more potently neutralizing FL-agnostic antibodies and broad and potent anti (α)- envelope dimer epitope (EDE) antibodies from the low-quality FL-sensitive antibodies. Further, analysis of the heavy chain complementarity determining region 3 (CDRH3) showed FL-sensitive and α-EDE antibodies were more negatively charged, but α-EDE antibodies were distinguished by longer and more aromatic CDRH3s. We then demonstrated how oligonucleotide-tagged DENV1-4 dimers and proteogenomic single-cell B cell receptor sequencing (LIBRA-seq) could identify memory B cells predicted to have potent neutralization based on DENV-binding and CDRH3 physiochemical properties. This study provides insight into the secondary DENV-specific B cell repertoire and approaches for identifying protective antibodies as part of large-screening efforts.

**One Sentence Summary:** Approaches to define the genetic, physicochemical, and functional properties of dengue virus envelope dimer-specific memory B cells after secondary infection.

## INTRODUCTION

Dengue viruses are the most prevalent flaviviruses globally and estimated to infect 390 million people annually (*1–4*). There are four antigenically distinct serotypes, dengue viruses 1-4 (DENV1-4), enabling reinfection in individuals who have been previously exposed (*5, 6*). Antigenic similarities between the serotypes contribute to disease observed in secondary DENV infection, where antibodies generated in the first infection have cross-reactive binding but minimal neutralizing activity to the new infecting serotype, facilitating viral replication through Fc receptor-mediated viral entry (*7–10*). This phenomenon, called antibody-dependent enhancement, is considered a driver of severe disease during secondary DENV infection and is an obstacle to developing safe and broadly protective dengue vaccines. A major target of enhancing antibodies is the conserved fusion loop (FL) region on the surface virus envelope (E) protein responsible for mediating viral and host cell membrane fusion during viral entry (*10–13*). In contrast, secondary infection with a heterotypic DENV serotype is thought to induce broadly neutralizing and protective antibodies such the anti (α)-E dimer epitope (EDE) and J8 and J9 antibodies, which target quaternary epitopes spanning E protein homodimers that cover the surface of the mature virion (*14–16*). However, the B cells encoding these antibodies have been found only in acute-phase plasmablasts. Thus, the memory B cells providing broad, long-lived protection are not well-characterized.

Our understanding of the B cell repertoire is hindered by limited tools for large-scale characterization of DENV-specificity and potency. Studies of DENV-specific memory B cells following secondary infection have often described their abundance and binding patterns using immunospot or selective enrichment, showing B cells collected long after secondary infection were more likely to bind four serotypes compared to those observed after primary infection (*17–20*). Studies of human monoclonal antibodies provide insight into specific epitopes and neutralization potency but only for a limited number of cells, and their contribution to the overall repertoire is not known (*18–23*). Most of these monoclonal antibodies bind to multiple serotypes and almost always neutralize the same serotypes to which binding is observed (*20, 23–25*). Antibodies targeting the FL are generally broad but more weakly neutralizing and include high avidity α-FL antibodies like 1C19 (*26*), which also bind the bc loop in E domain II (EDII) (*27, 28*). However, other cross-reactive epitopes are found in E domain III (EDIII) (*24, 29*). Interestingly, α-EDE antibodies target multiple of these epitopes, with a FL binding footprint in the heavy chain, and EDIII binding via light chain contact (*30*). However, no antibodies with potent and broad reactivity have been found in the human memory B cell repertoire.

Newer multiplexed, single-cell sequencing strategies like Linking B cell Receptor to Antigen specificity through sequencing (LIBRA-seq), in which panels of antigens are used to probe the specificity of individual memory B cells alongside their paired receptor sequences, present an elegant solution to characterizing these rare cell populations in resting individuals (*31, 32*). To take full advantage of these technologies, the flavivirus field needs reliable, sensitive probes that present quaternary, protective epitopes and can easily be labeled using fluorescent and oligonucleotide tags. Previous studies have used recombinant E proteins or fluorescently labeled virions as probes; however, recombinant E proteins primarily exist as monomers and do not present quaternary structures, and whole virion probes often require non-specific labeling that blocks epitopes and have high background staining due to non-specific binding to B cells (*22, 25, 33–37*). Stabilized E dimers with site-specific labeling sites are promising potential probes, as they enable detection of quaternary antibodies by presenting the E proteins in the dimeric conformation found on the mature DENV virion (*15, 30, 33, 38–40*). Further, as on the fully mature virion, the FL is tucked into the dimer interface, making it less accessible to antibodies than is found on E protein monomers or partially mature (‘standard’) DENV virions (*41–43*). However, even with reduced exposure of the FL, antibodies to flaviviruses are cross-reactive, and thus the interpretation of binding patterns from an assay in which B cells compete for antigens to the four serotypes is not obvious.

Here, we used an array of stabilized E dimers as next-generation probes for the detection and profiling of antigen-specific B cells by targeted proteomic and proteogenomic analyses. Through in-depth characterization of the DENV-memory B cell repertoire, we developed analytical approaches to interpret binding, genetic, and physicochemical features to aid in identifying memory B cells encoding broad, potently neutralizing antibodies.

## RESULTS

### DENV2-specific B cell isolation from an individual nearly three decades after last reported DENV infection

To characterize the repertoire of protective, long-lived memory responses of individuals years after DENV exposure, we recruited participants who had lived in dengue endemic areas and then resided for various lengths of time in the United States (NCT01306084). From this cohort, we selected a participant (DEN24) who reported two prior dengue cases, lived in an endemic area during a period of unusually high transmission, and received Japanese encephalitis virus (JEV) and yellow fever virus (YFV) vaccines. This participant had lived in the United States for nearly two decades prior to enrolling in our study, with minimal travel to endemic areas. Serum from DEN24 potently neutralized all four DENV serotypes when evaluated by standard (partially mature) virus neutralization assay and had similar neutralization titers against mature DENV2 and DENV3 but lower titers to mature DENV1 and DENV4 (Fig. 1A). DEN24 also had antibodies that neutralized other more distant flaviviruses (Fig. 1A), which identified them as a strong candidate to explore the cross-reactive memory B cell repertoire.

**Fig. 1.**
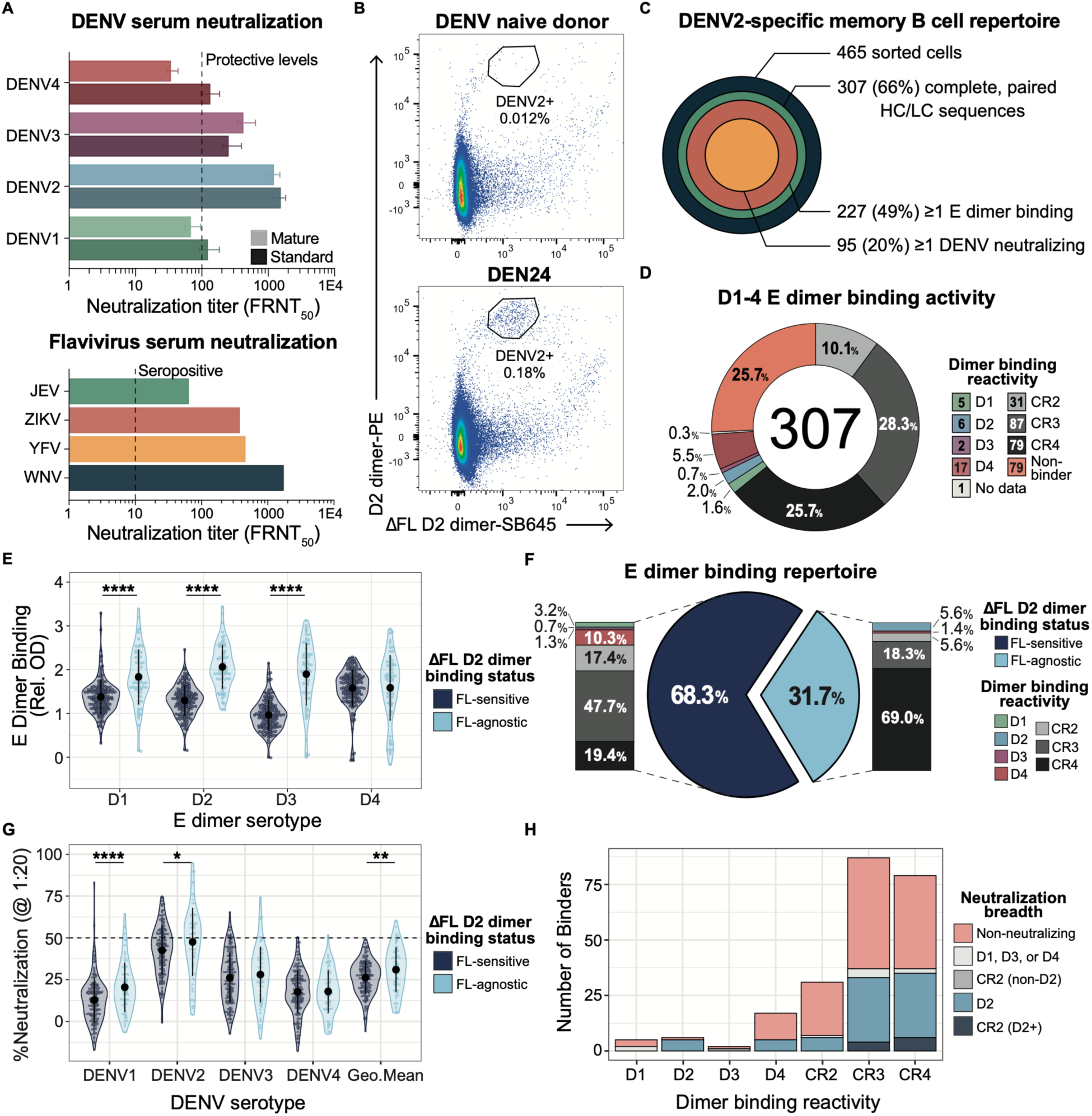
Characterization of DENV reactivity for antibodies isolated from DEN24 by RATP-Ig. **(A)** Standard and mature DENV1-4 and standard flavivirus (JEV, ZIKV, YFV, and WNV) serum neutralization titers (FRNT_50_) for DEN24 at the time of enrollment. Median FRNT_50_ titers from 18 independent standard virus and 6 mature DENV virus experiments are plotted in the bar graph, and error bars indicate the interquartile range. Protective mature DENV neutralization titer >100 is indicated as dashed line. **(B)** DENV2⁺ memory B cell frequency between DEN24 and a DENV-naïve donor, gated on the predominant antigen-specific population. **(C)** Bullseye summary of DEN24 screening for antibodies isolated by RATP-Ig. Concentric circle area is proportional to antibody number at each layer. **(D)** The 307 supernatants with paired heavy/light chain (HC/LC) sequences were screened for DENV reactivity and breadth by single-dilution ELISA against D1-4 dimers. Two supernatants lacked sufficient volume for assessment. **(E)** E dimer binding grouped by ΔFL D2 dimer binding status. Black points/lines indicate group mean ± standard deviation (SD). **(F)** Proportion of DEN24’s DENV-binding BCR repertoire classified as FL-sensitive (non-binding on ΔFL D2 dimer) or FL-agnostic (binding on ΔFL D2 dimer) and their corresponding E dimer reactivity breadths. **(G)** Percent neutralization of mature DENV1-4 at 1:20 dilution for 227 binding antibodies, grouped by ΔFL D2 dimer binding status. Black points/lines indicate group mean ± SD. Dashed line marks the 50% neutralization cutoff. **(H)** DENV neutralization breadth of the binding antibodies based on their dimer binding reactivity. **Abbreviations** – Geo. Mean: geometric mean; D1-4: D1-4 dimers; CR2-4: cross-reactive to 2-4 serotypes. Significance was evaluated by Wilcoxon rank-sum test. *p<0.05, **p<0.01, ***p<0.001, ****p<0.0001.

To define the DENV-specific memory B cell repertoire from DEN24 and confirm the utility of DENV E dimers as probes, we used the Rapid Assembly, Transfection, and Production of Immunoglobulins (RATP-Ig) pipeline. This approach enables simultaneous BCR sequencing and antibody production from single-cell sorted B cells through a series of immunoglobulin gene enrichment and nested PCR reactions designed to construct IgG1, κ transfection expression cassettes encoding extracted IgG and IgA V(D)J sequences (*44*). We piloted and refined the method using a donor with recent confirmed DENV infection (DEN20) (Fig. S1). For DEN24, we designed our antigen-specific cell sort to identify B cells expressing antibodies recognizing potent, quaternary epitopes using a stabilized DENV2 E dimer containing a single G106D substitution (ΔFL D2 dimer) that partially disrupts fusion loop-dependent binding of weak FL-‘sensitive’ antibodies (*39*). Given concern that the FL mutation may affect the orientation of the dimer and binding of quaternary epitopes, we also used the wildtype version of this dimer (D2 dimer). A site-specific biotinylation site was introduced into the C-terminus of each protein to enable unique fluorophore conjugation to the underside of the dimer, minimizing epitope occlusion. Magnetically isolated B cells were stained with the labeled probes (D2 dimer-PE and ΔFL D2 dimer-SB645) and then single-cell sorted using an inclusive gating strategy encompassing single- and double-positive D2 or ΔFL D2 dimer-specific IgD^−^ memory B cells (DENV2^+^ cells) with the gate set against cells from a DENV-naïve donor (Fig. S2). Restrictive gating of the apparent antigen-specific cell population identified a 15-fold greater frequency of DENV2^+^ cells for DEN24 than the DENV-naïve control (Fig. 1B).

### Functional screening of DENV2-specific B cells identified from DEN24

Of the 465 DENV2^+^ cells sorted using RATP-Ig, we recovered complete paired heavy and light chain sequences for 307 (66%) cells (Fig. 1C). They primarily expressed IgG receptors (93%), with a few cells encoding IgA (5%), while the remaining sequences lacked coverage to call isotype. We found 227/307 (74%) of the expressed supernatants from B cells with complete sequences bound to at least one of the DENV1-4 stabilized E dimers (D1-4 dimers), with 64.1% binding to two or more E dimer serotypes (CR2-4) by ELISA (Fig. 1D; Supplementary Methods). Only 9.8% bound one serotype, but many had binding to D2 just below our ELISA cutoff, suggesting they may also have been cross-reactive (Fig. 1D). We also measured antibody supernatant binding to the ΔFL D2 dimer (Fig. 1E-F). Antibodies that bound the D2 dimer but not the ΔFL D2 dimer interact at least partially with the FL and were classified as FL-sensitive due to their binding sensitivity at G106 in the FL. Such antibodies made up 68.3% (n=155) of the E dimer binding repertoire, demonstrating the immunodominance of the FL in this participant (Fig. 1F). Antibodies that bound both the D2 and ΔFL D2 dimers were classified as FL-agnostic antibodies, although they could interact with the FL at positions outside the mutated site. Notably, the FL-sensitive antibodies had lower binding absorbance to the D1-3 dimers compared to the FL-agnostic set (Fig. 1E) and only 19.4% bound all four serotypes (CR4), compared to FL-agnostic antibodies, where 69.0% bound four serotypes (Fig. 1F).

We then tested the supernatants for DENV neutralization by focus reduction neutralization assay (FRNT), defined as >50% neutralization at a 1:20 dilution of at least one mature DENV serotype (Supplementary Methods). Of the 227 dimer binding antibodies, 95 neutralized one or more serotypes (Fig. 1C). As observed for dimer binding, the FL-agnostic antibodies also had greater DENV1, DENV2, and geometric mean neutralization compared to FL-sensitive antibodies (Fig. 1G). Further, despite high cross-reactive binding, almost all neutralization was observed against DENV2, with 48% of FL-agnostic and 32% of the FL-sensitive antibodies neutralizing DENV2 (Fig. 1G). Even among the antibodies that bound three or four serotypes, the majority only neutralized DENV2, with a smaller fraction neutralizing both DENV2 and DENV3 (Fig. 1H). Thus, although most of the repertoire had cross-reactive binding to three or four serotypes, neutralization activity was only observed against one serotype.

### FL-agnostic antibodies have greater neutralization potency but restricted breadth

To further characterize the neutralizing memory B cell repertoire, we measured neutralizing antibody titers by 6-point FRNT against mature DENV1-4 for the top neutralizing antibodies to any serotype from the original screen with available binding data (Fig. S3A). Among this set, 57% bound to all four serotypes, and most only neutralized DENV2 and DENV3; however, they split evenly between FL-sensitive and FL-agnostic antibodies (Fig. 2A). From this screen, we selected 12 BCRs based on potent mature DENV2 and DENV3 neutralization and cross-reactive binding and produced purified monoclonal antibodies for further testing (Fig. S3B). Within this set, the FL-sensitive antibodies also had lower binding against the D1-3 dimers than the FL-agnostic antibodies (Fig. 2B), but there was a similar proportion that bound DENV4 between the groups (Fig. 2C). There were no differences in avidity (K*_D_*) for the wildtype D2 dimer between the FL-sensitive and FL-agnostic antibodies measured by biolayer interferometry (Fig. S3C), although the FL-agnostic antibodies had a significantly faster association rates (*k_on_*) than FL-sensitive antibodies (Fig. 2D), suggesting they had better access or binding to their epitopes on the E dimer.

**Fig. 2.**
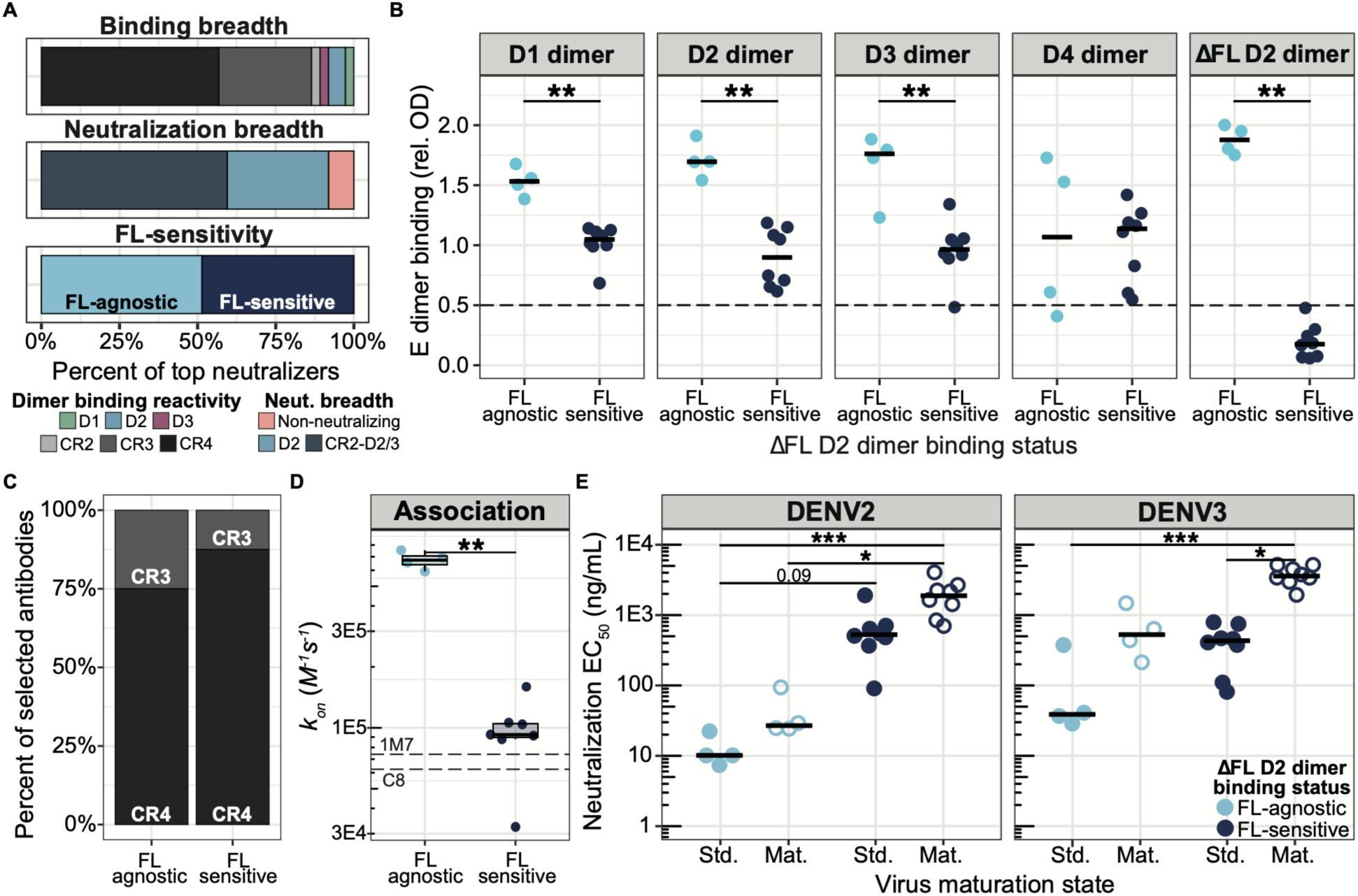
E dimer binding and flavivirus neutralization for FL-agnostic and FL-sensitive antibodies. **(A)** The E dimer binding breadth, DENV1-4 neutralization breadth, and FL-sensitivity of the top neutralizing antibodies (>60% neutralization to one or more DENV serotypes) during screening. **(B)** E dimer binding intensity of FL-agnostic and FL-sensitive purified monoclonal antibodies (rel. OD at 10 μg/mL) against D1-4 and ΔFL D2 dimers. **(C)** E dimer binding reactivities for the four FL-sensitive and eight FL-agnostic antibodies selected for further analyses. **(D)** *k_on_* rates (M⁻¹s⁻¹) for FL-agnostic and FL-sensitive antibodies, with *k_on_* rates for the known human antibodies FL-1M7 and EDE-C8 indicated by dashed lines. **(E)** DENV2 and DENV3 EC_50_ values for purified FL-agnostic and FL-sensitive antibodies determined by FRNT assay using standard and mature viruses. **Abbreviations** – EC_50_: half-maximal neutralization concentration; Std: standard virus; Mat: mature virus. Crossbars represent median values. Significance between groups was evaluated by Wilcoxon rank-sum test; between standard and mature EC_50_ values within/between groups by Kruskal-Wallis test with post-hoc Dunn’s test (Bonferroni-corrected). *p<0.05, **p<0.01, ***p<0.001.

However, the FL-agnostic and FL-sensitive antibodies differed significantly in their neutralization profiles. The FL-sensitive antibodies had broad neutralization: most antibodies neutralized standard DENV1-4, Zika virus (ZIKV), and a West Nile virus (WNV) vaccine candidate (WNV E and pre-membrane [prM] in an attenuated DENV4 backbone). However, neutralization breadth did not extend to the mature DENV viruses, with weak neutralization, which was only observed to mature DENV2 and DENV3 (Fig. 2E, Table 1, Fig. S4). In contrast, the four FL-agnostic antibodies potently neutralized both standard and mature DENV2 to similar or even greater degree than α-EDE antibodies and better than the FL-sensitive antibodies (Fig. 2E, Table 1, Fig. S4). Yet, although FL-agnostic antibodies potently neutralized standard DENV3, they lost neutralization to mature DENV3 and lacked the breadth to neutralize DENV1 and 4 like the α-EDE antibodies (Fig. 2E, Table 1). The only exception was low neutralization (EC_50_ = 6,799 ng/mL) of mature DENV1 observed for DEN24-4E01. Most FL-agnostic antibodies had minimal neutralization of non-DENV flaviviruses; only DEN24-5F07 neutralized standard ZIKV at an EC_50_ of 22 ng/mL (Table 1, Fig. S4). Based on cross-neutralization of DENV1-4 and ZIKV and low but detectable neutralization of mature virus (Fig. 2E, Table 1), the FL-sensitive antibodies resemble the high avidity α-FL antibodies described by others (*27*). The potent but restricted neutralization breadth of FL-agnostic antibodies suggested they may rely on serotype-specific residues for their neutralization activity.

**Table 1.** D2 E dimer binding avidity and flavivirus neutralization activity of top neutralizing antibodies^a^.

| Antibody | k<sub>on</sub><br>(x10<sup>5</sup>) | k<sub>off</sub><br>(x10<sup>-7</sup>) | K<sub>D</sub><br>(x10<sup>-12</sup>) | Standard DENV Neutralization |  |  |  |  | Mature DENV Neutralization |  |  |  |  | Flavivirus Neutralization |  |  |  |
| --- | --- | --- | --- | --- | --- | --- | --- | --- | --- | --- | --- | --- | --- | --- | --- | --- | --- |
|  |  |  |  | DENV1 | DENV2 | DENV3 | DENV4 | Geo Mean | DENV1 | DENV2 | DENV3 | DENV4 | Geo Mean | JEV | WNV | YFV | ZIKV |
| DEN24-4E01 | 6.9 | 5.2 | <1 | 7,571 | **10** | 375 | - | 733 | 6,799 | **25** | 636 | - | 1,017 | - | - | - | - |
| DEN24-5F07 | 7.5 | 155 | 20.5 | - | **22** | **41** | - | 550 | - | **94** | 1,476 | - | 1,930 | - | - | - | **22** |
| DEN24-5F06 | 5.9 | 3.4 | <1 | - | **7** | **29** | - | 381 | - | **29** | 213 | - | 888 | - | - | - | - |
| DEN24-5G02 | 6.6 | 169 | 25.8 | - | **10** | **37** | - | 439 | - | **24** | 438 | - | 1,015 | - | - | - | - |
| DEN24-2D08 | 0.9 | 4.8 | 5.2 | - | 1,911 | 468 | 8,814 | 2,980 | - | 2,243 | 3,393 | - | 5,253 | - | 186 | - | 1,274 |
| DEN24-3B04 | 1.6 | 4.1 | 2.6 | - | 483 | 750 | 3,067 | 1,826 | - | 4,040 | 4,491 | - | 6,527 | - | - | - | 7,923 |
| DEN24-1G02 | ND | ND | ND | 6,940 | 707 | 408 | 949 | 1,174 | - | 1,638 | 3,772 | - | 4,985 | - | 189 | - | 588 |
| DEN24-3E11 | 1.1 | 305 | 329 | - | **91** | **81** | 919 | 510 | - | 843 | 1,936 | - | 3,575 | - | 127 | - | 1,326 |
| DEN24-2E08 | 1.0 | 4.8 | 4.6 | 8,894 | 507 | 110 | 2,430 | 1,048 | - | 2,168 | 2,963 | - | 4,424 | - | 63 | - | 428 |
| DEN24-1F02 | 0.9 | 4.2 | 4.6 | 6,759 | 367 | 377 | 2,027 | 1,173 | - | 700 | 3,403 | - | 3,928 | - | 101 | - | 722 |
| DEN24-1A10 | 0.3 | 4.4 | 13.7 | - | 640 | 792 | - | 2,668 | - | 1,424 | 5,187 | - | 5,214 | - | 485 | - | 1,260 |
| DEN24-4C06 | 0.9 | 3.2 | 3.7 | - | 548 | 458 | - | 2,238 | - | 2,680 | 5,168 | - | 5,314 | - | 354 | - | 1,134 |
| EDE-C8 | 0.6 | 4.2 | 6.7 | 122 | **28** | 137 | 8,893 | 254 | 247 | **28** | 269 | 4,105 | 295 | - | - | - | 197 |
| EDE-C10 | 0.5 | 785 | 1700 | 246 | **26** | 218 | **83** | 104 | 396 | **31** | 1,023 | 118 | 197 | - | - | - | **44** |
| EDE-B7 | 1.5 | 817 | 532 | **30** | **45** | **71** | 217 | **68** | **42** | **26** | 110 | 614 | **93** | ND | ND | ND | ND |
| FL-1M7 | 0.7 | 31.3 | 42.2 | 158 | **48** | **57** | 398 | 115 | - | - | - | - | - | - | **62** | - | 171 |

<sup>a</sup>EC<sub>50</sub> values for standard and mature viruses (dash [-] indicates >10,000 ng/mL for DENV and >20,000 ng/mL for non-DENV flaviviruses; values <100 ng/mL in boldface). Color indicates FL-agnostic (light blue), FL-sensitive (navy), and control (grey) antibodies. ND: not determined.

### FL-agnostic antibodies map to EDII- and EDIII-specific epitopes

Given their distinctive properties and greater potency, we focused on the four FL-agnostic antibodies for epitope mapping by alanine-scanning prM/E mutagenesis libraries (*45*) (Fig. 3A-B). DEN24-4E01, DEN24-5G02, and DEN24-5F06 mapped to the A-strand of EDIII (Fig. 3A). For all three antibodies, residues E311 and I312 of the Α-strand were critical to binding (Fig. 3A). E311 is a highly conserved residue across all four DENV serotypes (Fig. 3C) and, among the sites we identified, was also an important EDIII residue for publicly available EDIII-specific antibodies (Fig. 3D). In contrast, I312 is restricted to DENV2 viruses and was not found in the epitope of other EDIII antibodies (Fig. 3C), which may help explain the potent neutralization of DENV2 observed for these antibodies. Unlike DEN24-5F06, whose binding centered on the Α-strand, DEN24-5G02 and DEN24-4E01 bound an additional residue within the G-strand at F392 (Fig. 3A). The F392 residue is conserved across DENV1, DENV2, and DENV4 (Fig. 3C), and is a key residue for ∼45% of known α-EDE antibodies (Fig. 3D), suggesting a contribution to broader binding.

**Fig. 3.**
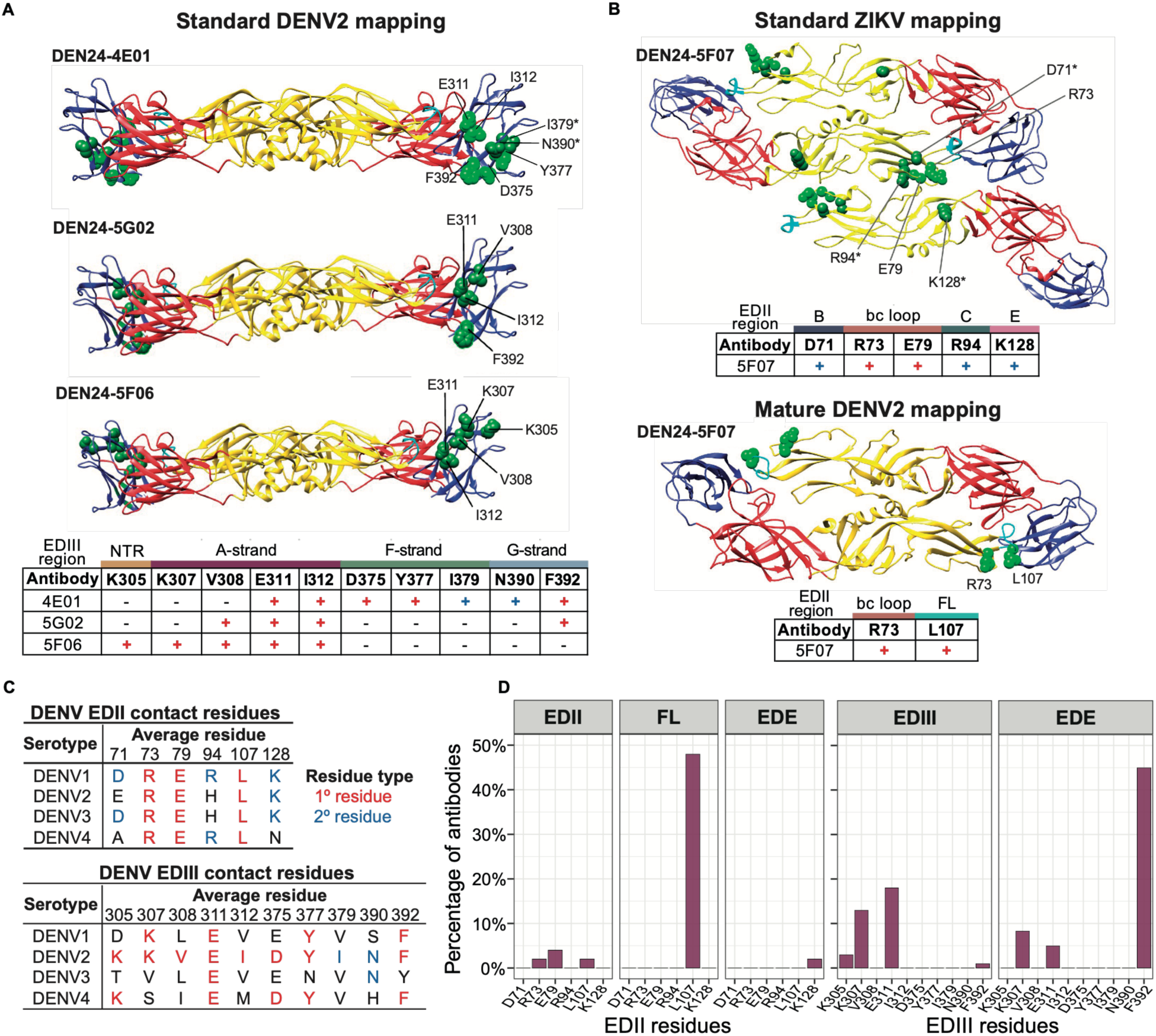
Epitope mapping of the FL-agnostic antibodies by alanine scanning shotgun mutagenesis library. Epitope mapping the four FL-agnostic antibodies to **(A)** EDIII determined by screening against the standard DENV2 prM/E alanine scanning library and **(B)** EDII based on the standard ZIKV prM/E or the mature DENV2 (prM/E co-expressed with furin protease) alanine scanning library. Ribbon diagrams indicating the structure of the D2 dimer (PDB 1OAN) with color coding for the domains of interest (EDI = red, EDII = yellow, EDIII = blue, and FL= cyan). Primary residues (red plus [+]), which contribute highly to the energetic favorability of binding, and secondary residues (blue plus [+]), which contribute moderately to binding, identified during screening are indicated in the table, while residues absent from an antibody’s epitope are indicated with a dash [–]. **(C)** The average amino acid residue identity for diverse DENV1-4 viruses included in Katzelnick *et al*. (*6*) for the EDII and EDIII contact residues identified for the four FL-agnostic antibodies. Amino acids are colored by their identity as primary (red) or secondary (blue) residues discovered during epitope mapping. **(D)** Frequency of residues identified that are present in publicly available α-EDII (n=46), α-FL (n=83), α-EDIII (n=90), and α-EDE antibodies (n=60) (*51*).

Ultimately, DEN24-5F07 mapped to the bc loop of E domain II (EDII, Fig. 3B). DEN24-5F07 failed mapping against the DENV2 prM/E construct, but since it also potently neutralized ZIKV, we mapped it against a ZIKV prM/E mutant library (*46*). ZIKV mapping identified R73 and E79 in the EDII-bc loop as primary critical residues for binding and D71, R94, and K128 as additional interacting secondary epitope residues (Fig. 3B, top). R73 and E79 are conserved residues within the EDII-bc loop for DENV1-4 (Fig. 3C) and ZIKV (*47*), which could explain the broad DENV binding and ZIKV neutralization observed. The epitope for DEN24-5F07 may be more complex than a standard EDII-bc loop antibody, as K128 can only be accessed as part of a quaternary binding site with the E-strand of an adjacent E dimer. Because this epitope is near the region of E blocked by prM, we also co-expressed the DENV2 prM/E library with furin protease to cleave pr and enable DENV2 mapping against mature E (Fig. 3B, bottom). This revealed a distinct footprint still centered on R73 but also included L107 in the FL, which is shared by nearly 50% of previously described α-FL antibodies (Fig. 3D), similar to the higher avidity FL-bc loop antibodies that arise after secondary DENV infection (*27*). However, the additional contact residues identified through ZIKV mapping may contribute to the lack of FL-sensitivity observed when screened against the ΔFL D2 E dimer. Together, these findings suggest that the binary categorization of low-quality α-FL antibodies versus highly neutralizing α-EDE antibodies only describes a fraction of cross-reactive antibodies, which instead exist along a spectrum of breadths, specificities, and potencies.

### Multiplexed E dimers identify distinct BCR binding patterns suggestive of neutralization activity

To determine whether antibody quality could be selected for based on E dimer binding profiles, we evaluated if B cell staining patterns from the RATP-Ig sort translated into antibody reactivities. We observed a weak negative correlation in D2 dimer and ΔFL D2 dimer fluorescence intensity across all the DENV2^+^ cells sorted (Fig. 4A), consistent with the D2 dimer binding preference observed even for the FL-agnostic antibodies by ELISA (Wilcoxon signed rank test, p<.001). However, there were still modest differences in the distribution of ΔFL D2 dimer binding B cells between the quadrant gates. For instance, the apparent antigen-specific population highlighted in Fig. 1B, marked by D2^hi^ΔFLD2^lo^ dimer staining (Fig. 4A), encoded mostly FL-sensitive antibodies (54% FL-sensitive, Fig. 4B). The small double-positive (D2^+^ΔFLD2^+^) population showed the greatest enrichment of FL-agnostic-encoding cells (40% FL-agnostic), while the population that favored the ΔD2 FL dimer (D2^lo^ΔFLD2^hi^) contained a higher fraction of non-binding cells (63% non-binders, Fig. 4B).

**Fig. 4.**
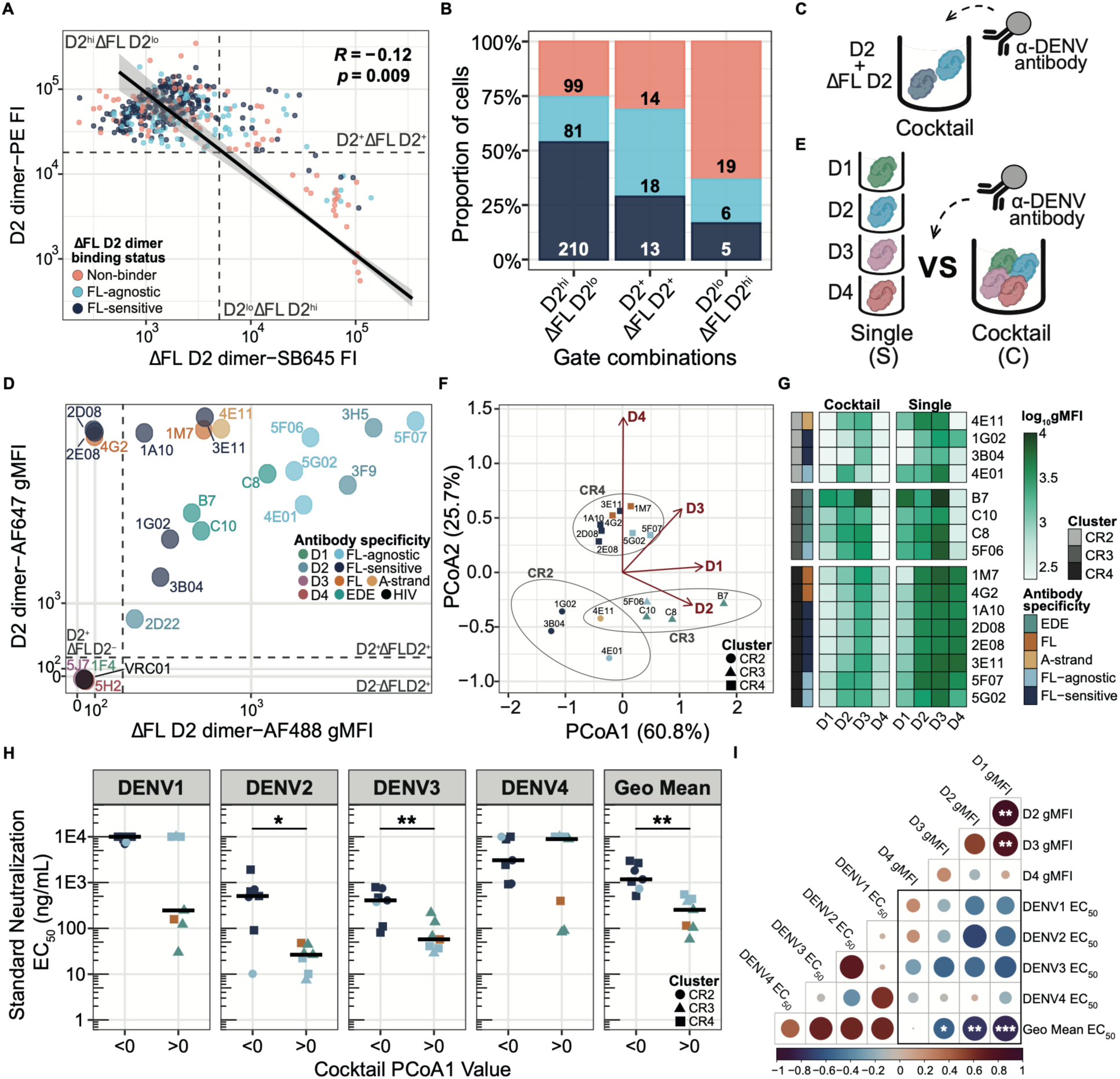
Preferential antibody binding profiles in a competitive system. **(A)** D2 and ΔFL D2 dimer fluorescent intensity (FI) for DENV2⁺ cells from DEN24’s sort, colored by ΔFL D2 binding status (Fig. 1F). Linear regression fit with Spearman *R* and p-value (top right). Quadrant gates for probe positivity shown as dashed lines. **(B)** Proportion of cells in each quadrant from A, encoding non-binding (orange), FL-sensitive (navy), or FL-agnostic (light blue) antibodies by ELISA. Cell counts per fraction is indicated. **(C)** Schema of probe cytometric bead assay: α-DENV antibody-coated beads were co-stained with D2 and ΔFL D2 dimers. **(D)** Geometric mean fluorescence intensity (gMFI) for D2/ΔFL D2 binding to beads labeled with cross-reactive α-FL, α-EDE, α-A-strand, α-D1-4 serotype-specific (TS), FL-agnostic, FL-sensitive, and α-HIV antibodies. Quadrant gates shown as dashed lines at 2×SD above the D2/ΔFL D2 gMFI for α-HIV antibody, VRC01. **(E)** Schema of probe competition for the cytometric bead assay, with single D1-4 dimer versus multiplexed D1-4 dimer cocktail staining of α-DENV antibody-coated beads. FL-sensitive antibodies DEN24-1F06 and DEN24-4C02 were not run. **(F)** Principal coordinates analysis (PCoA) performed using Manhattan distances calculated from standardized multiplexed D1-4 gMFI values. Antibodies are shaped by their k-means cluster assignment (CR2-4) and colored by antibody specificity. Fitted D1-4 vectors indicate the direction of increasing gMFI values. Antibodies positioned closer to, and in the direction of, a given probe vector exhibit relatively higher gMFI values to that probe. **(G)** D1-4 dimer gMFI values for cocktail- and single-stained beads labeled with cross-reactive and serotype-specific antibodies annotated by their antibody specificity and cluster identity from F. **(H)** Standard DENV neutralization EC_50_ values for antibodies with PCoA1 values <1 or >1 in F. Crossbars represent median values. Points are colored by their antibody specificity from in G and their shape denotes their cluster identity from F. Significance was evaluated by Wilcoxon rank-sum test. **(I)** Correlation matrix of multiplexed D1-4 gMFI values and DENV1-4 and geometric mean (Geo Mean) EC_50_ values. Dot size is proportional to the adjusted (Bonferroni-corrected) Spearman’s rank correlation coefficient, color gradation indicates directionality, and asterisks denote significant correlations. *p<0.05, **p<0.01, ***p<0.001.

To test if, among the most neutralizing antibodies, binding patterns to the two D2 dimer antigens could distinguish the highest quality antibodies, we used a cytometric bead assay that captured α-DENV antibodies on beads to mimic DENV-specific B cells (Fig. 4C). Co-staining with D2 and ΔFL D2 probes accurately identified serotype-specificity (e.g., 1F4 versus 3H5), FL-sensitive (e.g., 4G2), and FL-agnostic (e.g., 3H5) binding properties of known α-DENV antibodies, although staining was still greater for the wildtype dimer (Fig. 4D). However, cross-reactive antibodies demonstrated varied patterns of binding. The FL-agnostic antibodies had similar binding patterns to known DENV2-specific α-EDI antibody, 3F9 (*20*) and α-EDIII antibody, 3H5 (*48*), marked by bright staining for both probes (Fig. 4D). Conversely, the FL-sensitive and α-FL antibodies generally stained brightly for D2 with poor to no staining for ΔFL D2. However, the α-EDE antibodies had an intermediate staining profile for both dimers, suggesting weaker overall binding to DENV2 than the FL-agnostic antibodies (Fig. 4D). These data suggested D2 and ΔFL D2 staining alone could reliably identify FL-agnostic antibodies, but not separate α-EDE antibodies from FL-sensitive antibodies.

As α-EDE antibodies are known for their potent neutralization breadth, we suspected that BCR competition for D1-4 E dimer probes might reveal serotype binding preferences that would better distinguish potent versus weak neutralization activity. We measured binding of each antibody to D1-4 using a single- and multiplexed (cocktail)-stained version of the cytometric bead assay (Fig. 4E). Single-stained D1-4 binding patterns reproduced the cross-reactive binding patterns observed by ELISA (SFig. S5). However, the multiplexed-stained version showed distinct patterns. To interpret these data, we used principal coordinates analysis (PCoA), which treats the log-transformed, geometric mean fluorescence intensities (gMFI) of antibody binding to the four dimers as coordinates (Fig. 4F). Each dimer is represented as an axis in lower-dimensional space, and the position of the antibody along that axis interpretable as the magnitude of binding to that serotype relative to other antibodies. More balanced binding to D1-3 was identified by an antibody’s position along PCoA1, which accounted for 60.8% of the variance explained, whereas an antibody’s binding to D4 explained most of the position along PCoA2 (25.7% of variance, Fig. 4F). Using k-means clustering, the antibodies grouped based on low to moderate binding to D2 and D3 with minimal binding to D1 and D4 (CR2), balanced, strong binding to D1-3 with almost no binding to D4 (CR3), and strong binding to all four serotypes but with preferential binding to D3 (CR4, Fig. 4F-G). However, even within clusters, some antibodies had more balanced binding to D2 and D3, which placed them above zero along PCoA1. Overall, greater binding to D1-3 captured by PCoA1 > 0 was associated with more potent neutralization and being a FL-agnostic or α-EDE antibody (Fig. 4H-I). These findings suggest that methods incorporating BCR competition for antigens of multiple serotypes may reveal neutralization patterns and help discriminate higher-quality neutralizing antibodies from weaker, FL-sensitive antibodies.

### Distinct physicochemical, clonal, and germline signatures underlying fusion loop sensitivity

We next evaluated whether specific physicochemical properties of the complementarity determining region 3 (CDR3) could provide an alternative screening method for defining antibody potency and paratope specificity. Differences in CDR3 physicochemical properties did not differentiate neutralization capacity; however, CDRH3 did distinguish FL-sensitive versus FL-agnostic antibodies (Fig. 5A, Figs. S6-7). FL-agnostic antibodies had CDRH3s that were significantly less negatively charged (median 0 charge) than the FL-sensitive antibodies (median –1 charge, p<0.001), which complements the +0.93 charge estimated for the wildtype FL at neutral pH but not the ΔFL D2 dimer with a charge of –0.07 (Fig 5A, Fig. S8A). Additionally, acidic residue content and average polarity were significantly higher and the percentage of aromatic amino acid residues significantly lower for FL-sensitive CDRH3s than for FL-agnostic CDRH3s (Fig. 5A). The FL-sensitive repertoire also had significantly lower clonal diversity across diversity orders (*q*) compared to the FL-agnostic repertoire (Fig. 5B). Among the most expanded lineages (>4 clones), all contained both FL-sensitive and -agnostic antibodies. Yet within the largest clonal lineage, which contained six clones, the single FL-agnostic clone 2H07 had a more neutral charge than the four FL-sensitive antibodies (Fig. 5C), suggesting that these CDRH3 properties may also help select the best candidates within a lineage for further exploration.

**Fig. 5.**
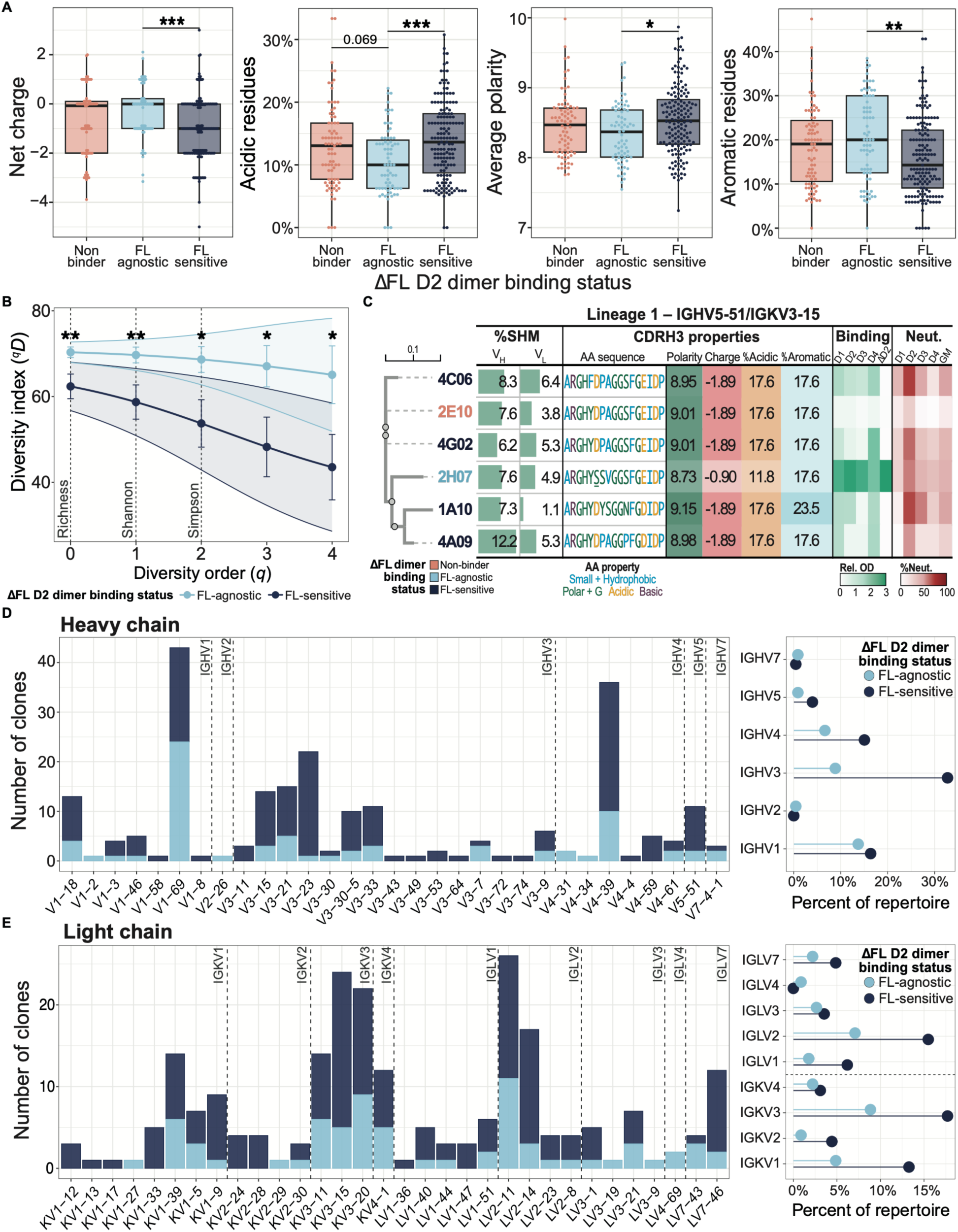
Physicochemical properties, clonal diversity, and germline usage associated with FL-sensitivity in DEN24’s DENV-memory B cell repertoire. **(A)** Physicochemical properties of the CDRH3 of isolated E dimer non-binding, FL-sensitive, or FL-agnostic antibodies. Significance by Kruskal-Wallis test with post-hoc Dunn’s test (Bonferroni-corrected). **(B)** Diversity indices (^q^D) of FL-agnostic and FL-sensitivity BCR sequences across continuous and fixed Hill number orders. Ribbons show 95% confidence intervals. **(C)** Neighbor-joining tree of Lineage 1 CDRH3 amino acid sequences (Clustal Omega alignment (*83*)) paired with a table of heavy/light chain variable region (V_H_/V_L_) somatic hypermutation rate (%SHM), CDRH3 physicochemical properties (sequence colored by property), and E dimer binding/DENV1-4 neutralization data. **(D)** Heavy chain variable and **(E)** light chain variable germline gene use across E dimer binding clones stratified by ΔFL D2 dimer binding status. To the right, the percentage of the total DENV E dimer binding repertoire attributed to each immunoglobulin gene family stratified by ΔFL D2 dimer binding status is shown. Abbreviations – GM: geometric mean; %Neut: %Neutralization at 1:20 dilution. *p<0.05, **p<0.01, ***p<0.001.

Germline usage was also associated with FL-sensitivity. DENV-reactive clones using *IGHV3* genes had the strongest bias towards FL-sensitivity, as 79% of *IGHV3* clones failed to bind ΔFL D2 dimer (Fig. 5D, right) and 95% of *IGHV3-23* clones were FL-sensitive (Fig. 5D, left). Most of *IGHV1* genes mapped to *IGHV1-69*-derived clones, which had a slight bias away from FL-sensitive binding (56% FL-agnostic) (Fig. 5D). *IGHV1-69* is a germline commonly used against glycosylated enveloped viruses, including DENV, and often produces broadly cross-reactive antibodies, most notably by the broadly neutralizing α-DENV antibodies J8 and J9 (*16, 49, 50*). Unlike *IGHV* gene use, patterns in FL-sensitivity for light chain genes depended more on the heavy chain gene to which it was paired (Fig. 5E). For example, most *IGVL2-11* clones paired with *IGHV1-69* were FL-agnostic, while *IGVL2-11* clones paired with *IGHV4-39* were predominately FL-sensitive (Fig. S8B).

### Distinct BCR features predict E domain epitope targets

Given these patterns, we were interested in whether the paratope features derived from sequences alone could be used to predict binding and epitope specificity. First, we used all CDRH3 properties and *IGHV* to predict FL-sensitivity using elastic net regression. In this model, charge and *IGHV3* use were the most important features, and aromaticity was moderately important (Fig. 6A). We then expanded our analysis to include an additional 173 publicly available human α-DENV CDRH3 sequences and their paired E epitope targets from a study that had evaluated BCR features associated with neutralization (*51*). They found polarity and aromaticity were important predictors of DENV neutralization (*51*). Here, we grouped our FL-sensitive antibodies with α-FL antibodies from the public sequences to determine if the same features could differentiate ‘FL-like’ antibodies from those targeting other domains. α-EDE antibodies contained CDRH3 properties distinct from antibodies targeting any other E domain epitopes and were particularly identifiable by longer and more aromatic CDRH3s (Fig. 6A-B). Conversely, low aromaticity was the most important feature for distinguishing FL-like antibodies. Furthermore, negatively charged CDRH3s were moderately important for predicting both FL-like and α-EDE antibodies. When we separately analyzed the CDRH3s of our FL-sensitive antibodies versus the previously published α-FL antibodies, those in our set were even less aromatic (Fig. 6B). These findings suggest the shared charge between FL-like and α-EDE antibodies may be due to their overlapping epitope footprints at the FL, whereas their contrasting aromaticity may contribute to their distinct neutralizing activities and breadths. Unlike the patterns observed for the overall binding repertoire of FL-sensitive antibodies, the median aromatic residue content of the neutralizing FL-sensitive antibodies described in Fig. 5 reached aromatic residue contents observed for α-EDE antibodies (Fig. 6C). This suggests high aromaticity, particularly at levels comparable to α-EDE antibodies, may be an important feature for distinguishing neutralizing FL-sensitive antibodies from their weakly or non-neutralizing counterparts.

**Fig. 6.**
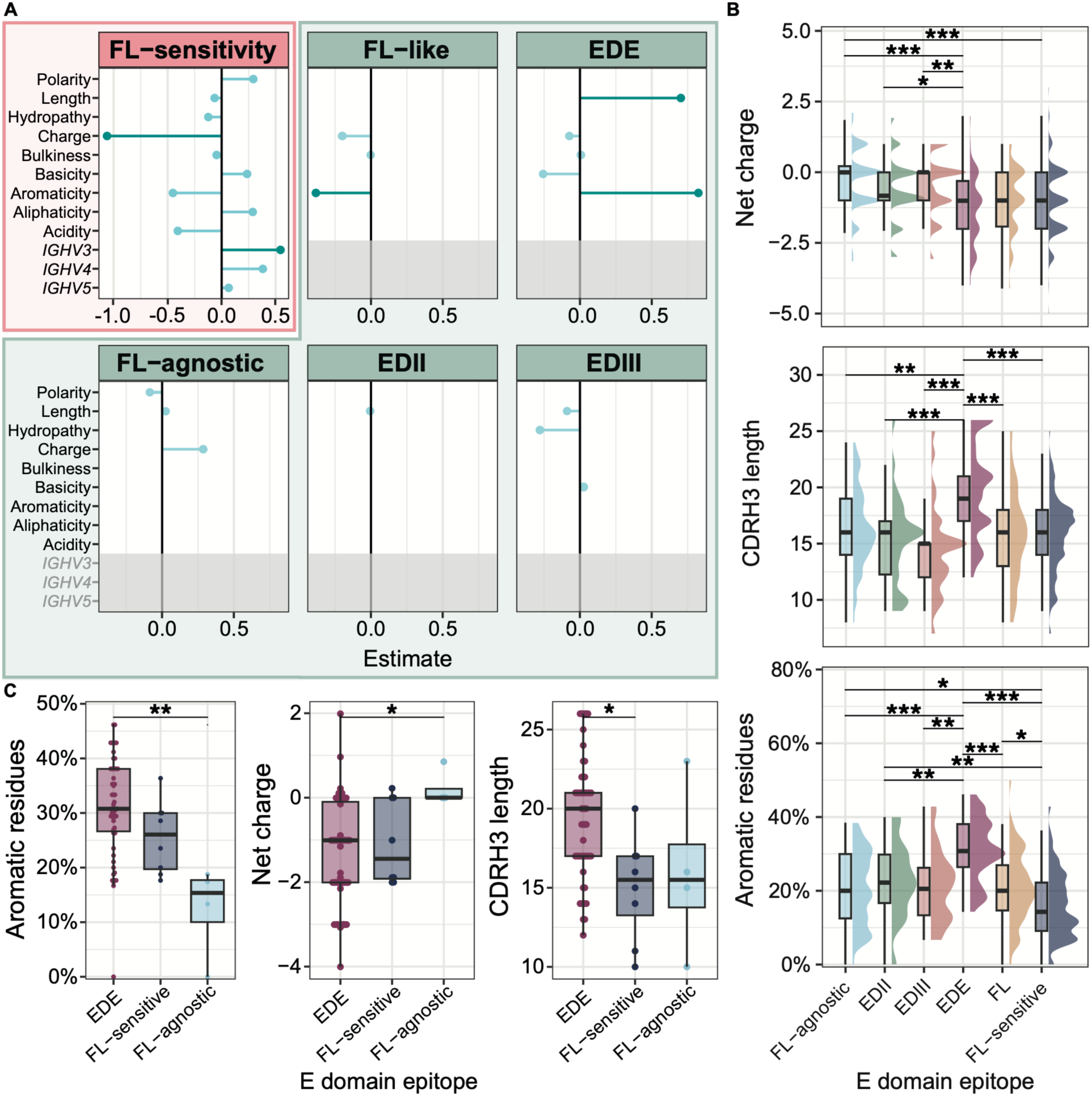
Important BCR features associated with FL-sensitivity and epitope targeting. **(A)** Estimated coefficients for CDRH3 physicochemical properties and *IGHV* gene family (*IGHV2*, *IGHV7* excluded; n≤3) from elastic net regression models predicting FL-sensitivity in DEN24 (top left, pink) or E domain epitope targeting in DEN24 combined with 173 publicly available human α-DENV antibodies (green) (*51*). Features with importance <40% are a lighter tint. Gene use was excluded from epitope predictions (grey region) due to unavailable germline data for public antibodies. FL-sensitive and public α-FL antibodies were combined for analysis (FL-like). **(B)** Important CDRH3 properties for DEN24’s DENV-specific memory B cell repertoire and publicly available human α-DENV antibodies (*27*), stratified by E domain epitope. **(C)** Important CDRH3 properties for publicly available α-EDE antibodies and the most potent neutralizing FL-sensitive and FL-agnostic antibodies from this study. Significance by Kruskal-Wallis test with post-hoc Dunn’s test (Bonferroni-corrected). *p<0.05, **p<0.01, ***p<0.001.

### Integrating E dimer binding profiles and BCR features for improved B cell screening

As functional screening of BCR cross-reactive binding profiles and CDRH3 physicochemical properties each improved screening for specific α-E antibody classes, we next sought to combine the two approaches into a single screening technique using proteogenomic single-cell BCR sequencing (LIBRA-seq) (*31*). While optimizing this method, we found staining of B cells with the E dimers labeled using oligonucleotide-tagged, fluorescent streptavidin was weaker than untagged, fluorescent streptavidin conjugates, and thus we used both oligo-tagged and untagged antigens in our sort (*52*). Here, we characterized 298 DENV-specific memory B cells identified using this approach from three donors. One donor had a prior primary DENV3 infection, and two had multiple prior DENV exposures, including a sample collected from DEN24 two years after the sample used in RATP-Ig. Shared clonotypes were identified between LIBRA-seq and RATP-Ig screens, including two antibodies from DEN24 with 100% homology for both the heavy and light chain CDR3 and similar binding profiles, and four more with >80% homology for the CDRH3 (Table S1). For the LIBRA-seq set, the majority of memory B cells had a strong preference for binding to a single serotype, and the cross-reactive cells almost all bound only two serotypes (Fig. 7A). For all three donors, memory B cell-specificities mirrored serum repertoires, favoring DENV2 and DENV3 reactivity, and the proportion of cross-reactive cells was associated with serum repertoire breadth (DEN24 > DEN12 > DEN1) (Fig. 7A). We also observed variation in DENV-specificity across B cell subsets between donors that may be explained by differences in DENV immunity or timing of sample collection relative to DENV exposure (Fig. 7B, Fig. S9, Table S2). For DEN12, the prominent DENV-specific population comprised of switched memory and exhausted B cells, especially for cells with greater binding breadths. However, we also observed modest type-specific or low cross-reactive DENV-staining by non-switched memory B cells and a few naïve B cells when we analyzed the total B cell compartment (Fig. 7B). We compared these results to DENV-reactivity across memory compartments measured by flow cytometry from a later sample from DEN12, demonstrating similar patterns to those observed by LIBRA-seq except for a loss in DENV-specific non-switched (IgM^+^) memory B cells by flow suggestive of waning (Fig. 7B).

**Fig. 7.**
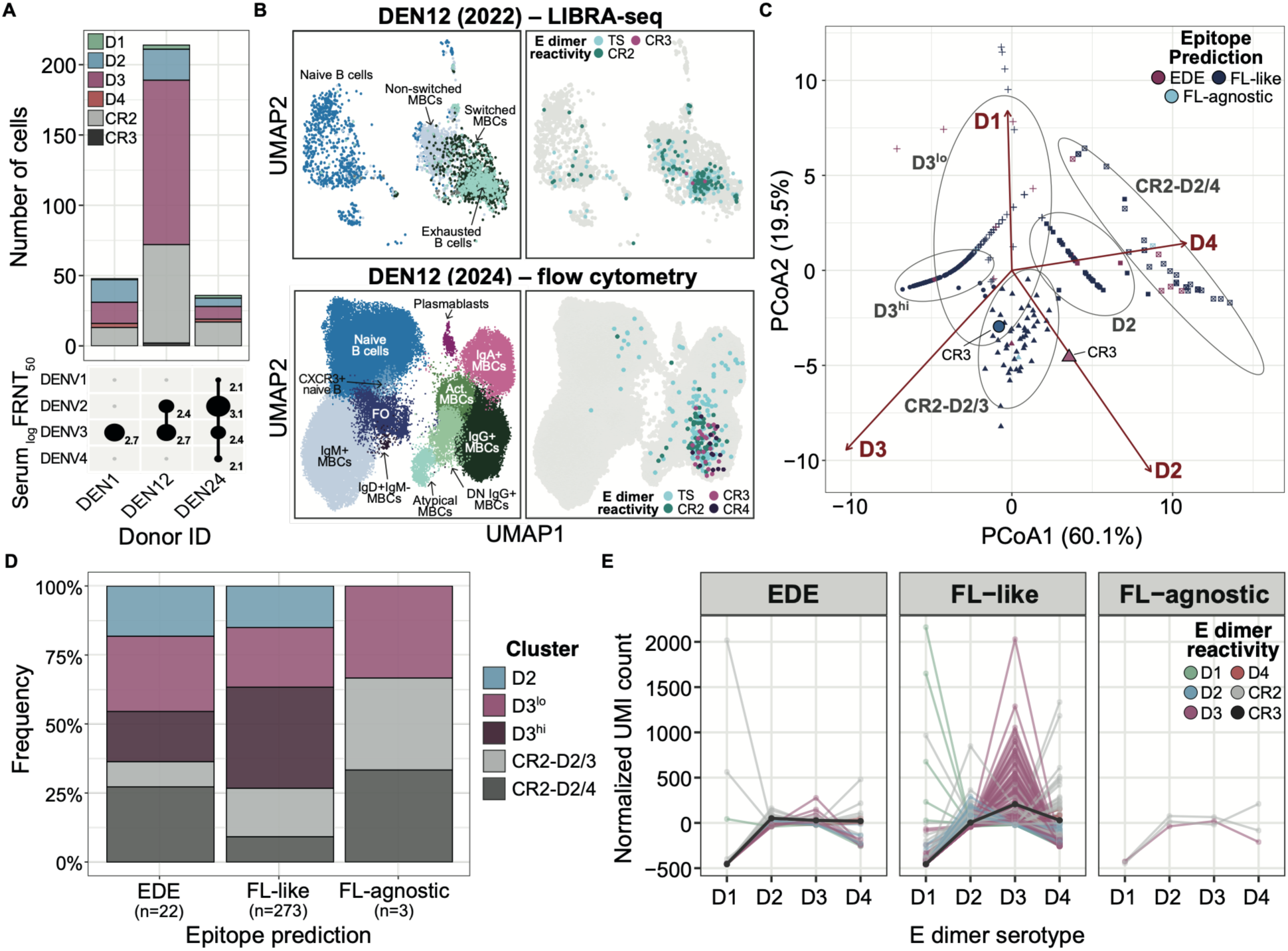
Identification, characterization, and predicted epitope of DENV-specific B cells using LIBRA-seq. **(A)** Number of DENV-specific memory B cells identified by LIBRA-seq, colored by E dimer reactivity. Circle size and values beneath the plot denote each donor’s standard DENV1-4 serum log_10_ FRNT_50_. **(B)** Uniform Manifold and Approximation Projections (UMAPs) of B cells from DEN12 collected 1.5 years apart, with each dot representing a cell, colored by cell subset annotation (left) and E dimer reactivity (right) as captured by LIBRA-seq (2022) and flow cytometry (2024). **(C)** PCoA performed using Manhattan distances calculated from normalized unique molecular identifier (UMI) values. Clones are shaped by their k-means cluster assignment (D3^lo^, D3^hi^, CR2-D2/3, CR2-D2/4) and colored by their predicted epitopes. Fitted D1-4 vectors indicate the direction of increasing UMI values. Clones positioned closer to, and in the direction of, a given probe vector exhibit relatively higher UMI values to that probe. **(D)** Proportion of clones in the assigned clusters from C for each predicted epitope with their respective cell count below. **(E)** D1-4 normalized UMI counts for clones assigned to each predicted epitope colored by their E dimer reactivity. **Abbreviations –** TS: type-specific binding to one serotype; D3^lo^: D3 binding preference with low UMI counts; D3^hi^: D3 binding preference with high UMI counts; CR2-D2/3: cross-reactive binding for D2 and D3; CR2-D2/4: cross-reactive binding for D2 and D4.

Applying the same unbiased clustering approach used to define DENV binding preference in the multiplexed-stained bead assay, the DENV-specific cells formed three serotype-dominant clusters (D2, D3^lo^, and D3^hi^) and two cross-reactive clusters characterized by D2 and D3 (CR2-D2/3) versus D2 and D4 (CR2-D2/4) binding (Fig 7C). We then used our physiochemical model to predict α-EDE, FL-like, and FL-agnostic BCRs based on CDRH3 sequences and compared how the cluster specificities were distributed across predicted epitopes. BCRs predicted to be EDE-like were enriched for cross-reactive clones (CR2-D2/3 and CR2-D2/4) (50%) compared to those predicted to be FL-like (32.6%) (Fig. 7D). The model only predicted three BCRs as FL-agnostic, with two found in the cross-reactive clusters. In contrast, the FL-like BCRs mapped predominantly to the type-specific clusters, with a majority exhibiting dominant D3 binding. This was consistent with the D3 dominance observed during the competition bead assay for the known α-FL and FL-sensitive antibodies isolated from DEN24 by RATP-Ig (Fig 4F-G). Thus, we observed greater cross-reactivity for the predicted EDE-like antibodies and more serotype-dominance in the FL-like antibodies, consistent with the bead assay results.

Only two BCRs were defined as having cross-reactive binding to three serotypes based on our cutoffs (Fig. 7C). One was predicted to be FL-like and exhibited a strong D3 binding preference (D3^hi^ cluster, Fig. 7C&E). Derived from an *IGHV3*-expressing non-switched memory B cell, it shared similar properties as our FL-sensitive antibodies from the RATP-Ig screen. In contrast, the second BCR was predicted to be EDE-like and had more balanced D2-4 binding (Fig. 7E). This BCR featured an *IGHV1-69*06/IGHJ6*03* heavy chain rearrangement and was expressed on a switched memory B cell, sharing key properties with previously described potent α-DENV antibodies (*16, 53*).

## DISCUSSION

Here, we used complementary sequencing approaches to show how multiplexed E dimer antigen binding profiles and BCR sequence features can be used to define the DENV immune repertoire and identify more potently neutralizing antibodies. We confirmed that a high proportion of antibodies derived from probe-positive cells bound to D1-4 E dimers by ELISA, and 42% of those that bound DENV were also neutralizing. In-depth characterization of the binding and neutralizing memory B cell repertoire in a highly exposed donor identified a mixture of FL-sensitive and FL-agnostic antibodies with broad binding but neutralization mostly limited to DENV2 and DENV3. Among the top neutralizers, antibodies that bound the FL had greater neutralization breadth, whereas those targeting other epitopes had more potent, specific neutralization. We also identified CDRH3 features that predicted epitopes: negative charge was associated with contacting the FL, but longer CDRH3s and greater aromaticity distinguished α-EDE antibodies from other FL-targeting antibodies. In addition, we confirmed multiplexed D1-4 dimers could identify type-specific and cross-reactive binding patterns by both targeted proteomic and proteogenomic analyses, demonstrating the applications for which E dimer probes can provide additional insight in characterization of DENV humoral responses and B cell memory.

A majority of DENV E dimer-specific memory B cells isolated from a highly exposed individual had BCRs with cross-reactive binding to three or four serotypes but neutralization of only one or at most two serotypes. FL-sensitive antibodies were dominant (68%), and clonal enrichment was observed primarily in the FL-sensitive repertoire. Prior work has suggesting greater clonal diversity emerged in the absence of the immunodominant FL, where vaccination of mice with DENV2 E monomer induced a network comprised of a single, highly connected clone with numerous disconnected clones, whereas mice receiving DENV2 EDIII or ZIKV E/prM with mutated FLs developed multiple major clonotypes (*53*) and had more diverse V-gene use in germinal center B cells (*54, 55*). We also observed a strong bias towards FL-sensitivity by *IGHV3-23*-encoding cells, which were enriched in DENV, ZIKV, and YFV-infected and -vaccinated individuals (*36, 56–62*), suggesting that one of the most abundantly used *IGHV* genes in the human BCR repertoire efficiently produces α-FL antibodies (*55*). Within the overall repertoire, the FL-sensitive antibodies were cross-reactive binders but bound fewer serotypes and had less neutralization than FL-agnostic antibodies. However, among the neutralizing, cross-binding antibodies, a similar number were FL-sensitive and FL-agnostic, and most neutralized both DENV2 and DENV3. The best FL-sensitive antibodies had broader neutralization, even neutralizing non-DENV flaviviruses, but were much weaker neutralizers compared to FL-agnostic antibodies. However, given their ability to neutralize mature DENV, these FL-sensitive antibodies are likely higher avidity α-FL antibodies that arise after secondary infection and not the classic, weakly cross-reactive α-FL antibodies that contribute to enhancement (*27*). The broad, strong binding observed for FL-agnostic antibodies contrasted with their limited neutralization breadth, likely because they targeted a mix of conserved and DENV2-specific residues. Prior work has identified subcomplex antibodies targeting EDIII, but the serotypes to which binding was observed were also the serotypes that were neutralized (*24, 29*). The observation that a large fraction of the post-secondary immune repertoire is broadly binding but narrowly and potently neutralizing points to an underappreciated class of antibodies.

Memory-derived BCRs are believed to favor breadth over affinity to recognize variant antigens and quickly aid in viral clearance and B cell maturation (*63*). Memory B cells encoding cross-binding antibodies, either FL-agnostic or FL-sensitive, may help protect highly exposed individuals against serotypes to which they have sub-protective serum antibody titers, efficiently reducing viral load and disease progression. The FL-sensitive and FL-agnostic antibodies each have some of the properties of α-EDE antibodies, and perhaps in combination provide similar protection. α-EDE antibodies have heavy chains centered on the FL and light chains that bind the A-strand of EDIII in α-EDE1 antibodies and the N153 glycan on EDI in α-EDE2 antibodies (*30*). The contact residues for α-FL antibodies also map to the heavy chain, like α-EDE antibodies, suggesting that improved light chain engagement of complex epitopes on adjacent E domains may be a mechanism by which α-FL antibodies can gain α-EDE antibody activity (*64*). The most potent FL-agnostic antibodies we identified also bound the EDIII A-strand epitope but lacked the contact in the FL like α-EDE antibodies.

Incorporating multiplexing strategies into repertoire characterization and antibody screening can simultaneously improve throughput and expand the scope of data collected. However, we also demonstrated that multiplexing E dimer probes during staining created a competitive environment that captured the intrinsic serotype binding preferences of cross-reactive B cells and antibodies. For our most neutralizing antibodies, their competitive D1-4 binding profiles were associated with their neutralization potency, with serotype-biased binding observed for the less potently neutralizing α-FL and FL-sensitive antibodies and more balanced binding observed for potently neutralizing α-EDE and FL-agnostic antibodies. In fact, preferential binding to one serotype was sufficient to identify FL-sensitive antibodies without the use of the ΔFL D2 dimer and was a profile shared by the majority of predicted FL-like cells in our LIBRA-seq data. This underlying serotype preference observed in the competitive assay for FL-like antibodies generally mirrored the serotype dominance observed for DENV1-4 binding avidities of the α-FL antibodies 1M7 and 4G2 measured against virus-like particles by biolayer interferometry (*65*), which may explain why α-FL antibodies enhance some serotypes but are more neutralizing against others. Prior studies have exploited the inherent competitive environment of LIBRA-seq to design formal competition-driven screens, such as target-ligand blocking to screen for neutralizing antibodies by selecting clones that blocked SARS-CoV-2 spike probes from binding to a soluble version of their receptor, angiostatin-converting enzyme 2 (*66*). This suggests that BCR binding profiles determined by multiplexed probes are partly a measure of avidity from the competitive nature of the antigen stain and thus may provide more nuanced information on specificity and potency.

Consistent with CDRH3’s critical role in determining paratopes (*67*), we found differences in CDRH3 sequences associated with antibody specificity. The most important predictor of FL-sensitive antibodies was CDRH3 charge. Long-range electrostatic interactions are important for directing proteins to regions of complementary charge (*68*), and the loss of charge in the ΔFL D2 dimer may be a primary mechanism underlying the FL-sensitivity observed. Antibodies initially form weakly specific, charge-based complexes with antigen, followed by precise docking mediated by short-range interactions to form a higher affinity complex (*69*). While more negatively charged CDRH3s were also important for predicting α-EDE antibodies, the most prominent predictors were long CDRH3s (median 20 amino acids) and high aromatic residue content (median 31%). Both features could facilitate more specific docking after initial contact through improving binding of the E protein surface and its glycans (*30*). Extra CDRH3 length helps the loop slip past glycans to enable direct contact with the peptide (*70*), and aromatic rings encourage CH-*π* interactions at the paratope-epitope interface as well as with glycans (*71*). A recent study using sequence-based modeling and molecular docking showed greater polarity and aromaticity were critical properties for predicting DENV-neutralization antibodies, mediating binding by maintaining the proper secondary structure of paratope-epitope interaction (*51*). While the overall FL-sensitive population identified here had very low aromaticity, the potently neutralizing FL-sensitive antibodies had aromaticity levels similar to α-EDE antibodies. These neutralizing FL-sensitive antibodies may thus represent an intermediate paratope composition between enhancing α-FL and protective α-EDE antibodies that is enriched for CDRH3 properties associated with better DENV neutralization. However, these FL-sensitive antibodies may still lack other key properties of α-EDE antibodies that provide their signature broad neutralization potency.

Our study has several limitations. The FL-sensitive signature identified here was restricted to α-FL antibodies sensitive to the G106 mutation in the FL. Further, the in-depth repertoire characterization was limited to one donor and may reflect DEN24’s unique immunity, infection sequence, and host genetics. Their history of DENV infection followed by flavivirus vaccination may also be different from a highly exposed individual whose most recent exposure was to DENV. While that makes it difficult to assert whether the prevalence and predictors of FL-sensitive antibodies observed here is representative of the DENV-memory B cell repertoire of highly exposed individuals, we identified similarities in CDRH3 properties between publicly available α-FL antibodies and our FL-sensitive antibodies as well as in two other donors in the LIBRA-seq analysis. An additional limitation is that the validation of the D1-4 dimer probes on primary B cells was performed on a small number of individuals. However, we did use the same individuals between methods, which enabled comparison of different approaches.

Generating B cell responses that overcome antibody-dependent enhancement driven by cross-reactive antibodies has been the greatest challenge in developing safe and effective dengue vaccines. Integrating advanced experimental and computational approaches to uncover features of protection within the α-DENV B cell repertoire will be essential to resolving the humoral complexity of α-DENV immunity and to moving the field closer to durable, broadly protective interventions. We demonstrated above that an integrated screening approach combining functional screening and BCR feature prediction led to the identification of a broadly competitive, cross-binding B cell with signature features of α-EDE antibodies including the use of *IGHV1-69*, which can produce broadly cross-binding, FL-agnostic antibodies, and was also used by previously described broadly neutralizing α-DENV antibodies (*16*). The antibody specificity screening and CDRH3 signatures described herein can be used in future repertoire and antibody discovery studies to guide computational pre-screening and further facilitate functional screening for more potently neutralizing antibodies. Further, the paired functional and physicochemical context regarding paratope-epitope properties of α-DENV binding and neutralizing antibodies further informs rational vaccine design and therapeutic monoclonal development. Finally, the adaptation of stabilized E dimers for cellular characterization pushes the field toward an integrated approach to study the diversity and targets of the protective versus enhancing antibody repertoires alongside the development of the cells encoding them.

## MATERIALS AND METHODS

### Study Design

Ethical approval was obtained from the Institutional Review Board of the National Institutes of Health and is registered at clinicaltrials.gov (NCT01306084). Written informed consent was obtained from participants prior to enrollment. Upon enrollment, participants completed a survey that provided demographic information (e.g., age, sex, ethnicity, city of birth, years living in and outside endemic dengue regions) and details on prior DENV infections, risk factors, and flavivirus vaccination history. Peripheral blood and serum were collected by venipuncture. Serum from the time of collection was used to characterize DENV immune status by a standard neutralization assay. Peripheral blood, collected in Acid Citrate Dextrose Vacutainer™ collection tubes (BD Biosciences), was centrifugated to first remove plasma, which was clarified and frozen at -80°C. PBMCs were then isolated by density gradient centrifugation using SEPMATE™-50 (STEMCELL Technologies) tubes per the manufacturer’s instructions. Residual red blood cells were lysed using Red Blood Cell Lysis Buffer (Sigma-Aldrich) or TheraPEAK® ACK Lysing Buffer (Lonza) and counted using a hemocytometer or Guava® easyCyte™ flow cytometer (Cytek). PBMCs were cryopreserved in 90% heat-inactivated fetal bovine serum (FBS) and 10% dimethyl sulfoxide for 24h at -80°C before transferring to -140°C for storage. Serum was collected using SST tubes (Vacuette) per manufacturer’s recommendation and stored at -80°C.

### Antigens

The DENV1-4 stabilized E dimers (D1-4) (*39, 40*) and the ΔFL D2 E dimer containing a G106D mutation to disrupt fusion loop binding (*39*) were expressed and purified by the UNC Protein Expression and Purification Core using the following, previously published, construct designs and expression plasmids: DENV1 (I2-U6-P4-S1-Avi), DENV2 (EV8 I2-U6 and EV8 I2-U6-Avi), DENV3 (I9-U6-P4-S1-Avi), DENV4 (I2-U6-P4-S1-Avi), and ΔFL DENV2 (I2-I8-U6 and I2-I8-U6-Avi). Constructs were modified to include an AviTag sequence (Avi) on the C-terminus of the protein to enable site-specific biotinylation. 100 μg of mammalian BirA ligase expression plasmid was co-transfected with the expression plasmid DNA in media supplemented with 100 μM biotin during expression of AviTag-containing constructs to generate biotinylated proteins. Proteins were stored at -80°C until use, and thawed proteins were held in 4°C storage to avoid freeze-thaw cycling. Commercial biotinylated SARS-CoV-2 spike proteins from strains D614G (Acro Biosystems, SPN-C82E3) and BA.1 (Acro Biosystems, SPN-C82Ee) and Human Serum Albumin protein (Acro Biosystems, HSA-H82E3) were frozen in small aliquots and thawed fresh each use.

### Fluorescent Streptavidin Conjugation of Antigen Probes

Biotinylated proteins were labeled for use as antigen-specific B cell probes using fluorescently labeled streptavidin. For the DENV-specific sort of DEN24 for RATP-Ig, 5 μg of D2 and ΔFL D2 E dimer proteins were combined with 2 μg of streptavidin-PE (12-4317-87, eBioscience) or streptavidin-SB645 (64-4317-82, eBioscience) respectively with PBS to total 30 μL and incubated overnight at 4°C. A staining concentration of 2.5 μg/mL of labeled antigen was used. All other experiments used the following conjugation strategy. Biotinylated D1-4 dimers, SARS-CoV-2 (CoV2) spike, and HSA proteins were conjugated with fluorescently labeled streptavidin each morning prior to use using the following procedure. Biotinylated protein and fluorescently labeled streptavidin were combined in a 1.23 μM ratio of DENV antigen:streptavidin or 0.75 μM ratio of SARS-CoV-2/HSA antigen:streptavidin in PBS. Optimal antigen:streptavidin ratios were determined during optimization experiments conducted previously (*52*). See Table S2 for antigen-fluorophore pairings. TotalSeq-C oligonucleotide conjugated streptavidin (BioLegend, 405283, 405285, 405293, 405159, 405195, 405197, 405199) was used to label antigens for LIBRA-seq. Antigen-streptavidin assemblies were incubated for 1h at 4°C. To block unbound streptavidin before use, antigen assemblies were quenched with 200 μM D-biotin (Invitrogen, B20656) for 30 min and centrifuged at 2,500xg for 5 min to pellet large aggregates. During antigen-specific staining steps, 1.65 μL of labeled antigens (5 μg/mL) were added to corresponding samples in a final staining volume of 50 μL or pooled together into an antigen cocktail for multiplexed staining.

### DENV-Specific B Cell Sorting using D2 E Dimer Proteins

Cryopreserved PBMCs from a DENV-naïve and the DENV-immune donor, DEN24, were thawed, counted, and B cells were isolated from PBMCs using the EasySep™ Human B Cell Isolation Kit (STEMCELL Technologies) per the manufacturer’s recommendation. Isolated B cells were stained with D2 dimer probes, a B cell phenotyping panel, and a viability dye to exclude dead cells. Cells were treated with Human Fc Block (BD Biosciences) for 10 min prior to antibody staining. After Fc blocking, cells were stained with 14 μg/mL of pre-conjugated D2-PE and ΔFL D2-SB645 probes for 30 min before staining with LIVE/DEAD Fixable Blue Dead Cell Stain (1:32 dilution) (Invitrogen) and the following B cell phenotyping panel for 20 min: 1:13 dilution of CD19-BB700 (HIB19, BD Biosceinces), 1:13 dilution of CD19-BUV563 (SJ25C1, BD Biosciences), 1:13 dilution of CD27-BV711 (O322, BioLegend), 1:53 dilution of IgD-BV786 (1A6-2, BioLegend), 1:53 dilution of CD21-AF700 (Bu32, BioLegend), 1:13 dilution of CD72-FITC (3F3, BioLegend), 1:6 dilution of FcRL4-PE/Cy7 (413D12, BioLegend), 1:6 dilution of FcRL5-APC (509f6, BioLegend), 1:6 dilution of IgK-APCFire570 (MHK-49, BioLegend), 1:53 dilution of CD85j-BUV737 (GHI/75, eBioscience), 1:24 dilution of CD32-SV500 (FUN-2, BioLegend), and 1:13 dilution of CD11c-BV421 (S-HCL-3, BioLegend). Dead cells were excluded and IgD^−^ memory B cells specific for the E dimer(s) (DENV2^+^) were sorted using gates set on a DENV naive donor (see Fig. S2 for sorting strategy). All DENV2^+^ B cells were single-cell sorted into 96-well plates containing 5 μL of TCL (Qiagen) buffer with 1% β-mercaptoethanol using a BD FACSymphony S6 and frozen at -80°C to be preserved for RATP-Ig high throughput monoclonal antibody production and repertoire characterization (*44*).

### Rapid Assembly, Transfection, and Production of Immunoglobulins (RATP-Ig)

Sorted DENV2^+^ memory B cells frozen in lysis buffer underwent BCR sequence isolation and expression cassette synthesis using a method called RATP-Ig described previously (*44*). Briefly, cDNA was synthesized from single-cell RNA extracts using a modified 5’RACE (rapid amplification of cDNA ends) protocol, which underwent PCR enrichment for heavy and light chain immunoglobulin variable regions. A portion of this product was reserved for Nextera library preparation using Unique Dual Indexes (Illumina) and paired-end sequencing (2×150bp read length) using an Illumina MiSeq. A separate aliquot was used to synthesize and assemble expression cassettes containing heavy and light chain fragments that were co-transfected for monoclonal antibody production. Assembled cassettes were amplified and transfected into Expi293 cells in 96-well deep-well plates using the Expi293 Expression System (ThermoFisher Scientific) per the manufacturer’s instructions. Following 5-7 days of incubation at 37°C, transfection supernatants were harvested, clarified, and held at 4°C throughout screening or stored in -80°C for long-term storage. Purified monoclonal antibodies were made from V(D)J sequences recovered during BCR sequencing and produced as IgG1, κ antibodies using GenScript’s high throughput antibody production service.

### RATP-Ig BCR Sequencing and CDR3 Analyses

V(D)J sequence analysis was performed as previously described for RATP-Ig (*44*). Illumina reads were demultiplexed, followed by paired heavy and light chain reconstruction using BALDR (*72*). Outputs were filtered to remove contigs with low coverage and/or incomplete sequences based on nucleotide length or missing J genes using filterBALDR (https://github.com/scharch/filterBALDR). Passing sequences were annotated using the single-cell mode in SONAR v4.2 (*73*) using IMGT gene nomenclature (*74*). Clonotypes were defined as cells with matching V genes and 80% nucleotide similarity for both heavy and light chain CDR3s. Non-productive rearrangements and any cells that 1) mapped to more than one productive heavy and light chain or 2) were missing a paired heavy or light chain sequence were excluded. Physicochemical amino acid property predictions for antibody CDR3 regions and clonal diversity analyses were performed using the alakazam package in R (*75*). Clonal diversity (*D*) was calculated as a continuous function of *q*, ranging from 0 to 4 at intervals of 0.1.

### Protein G Antibody Capture Cytometric Bead Assay

A cytometric bead assay designed to capture known α-DENV monoclonal antibodies and detect DENV E dimer binding profiles was adapted as described previously (*52*). Serotype-specific (1F4 (*22*), 3H5 (*48*), 2D22 (*22*), 3F9 (*20*), 5J7 (*76*), and 5H2 (*77*)) and cross-reactive (4E11 (*78*), 1M7 (*20*), 4G2 (*48*), EDE-C8 (*15*), EDE-C10 (*15*), and EDE-B7 (*15*)) α-DENV antibodies, as well as the α-HIV antibody VRC01 (*79*), were captured on Dynabeads™ Protein G magnetic beads (Invitogen) and stained for 30 min at 4°C with 10 μg/mL of with pre-conjugated fluorescent D1-4/ΔFL D2 dimer probe (single-stained) or a cocktail containing either D2-AF647 and ΔFL D2-AF488 or D1-4 dimers (multiplexed) (Table S3). Samples were fixed and acquired next day on a Sony ID7000 spectral analyzer. Data were analyzed using FlowJo 10.10.0 software (BD Biosciences).

### LIBRA-seq

Antigen-specific B cells sorted from one DENV-naïve and three DENV-immune donors were single cell sequenced in a previous report (data and analyses available on Github: https://github.com/foocheung/23_303) (*52*). Dual labeling of cells with antigen conjugated to generic streptavidin and oligonucleotide-tagged streptavidin were used to improve sensitivity using the following antigen panel: D1-4 E dimers, D614G and BA.1 SARS-CoV-2 spikes and HSA. BCR sequences and cell annotations were extracted from sample metadata files for cells from DENV-exposed individuals. DENV-positivity was defined using UMI count threshold of > 25 in the antigen (DENV1-4 E dimers) condition and <25 in the negative controls (D614G and BA.1 SARS-CoV-2 Spike and HSA). Subsequently, a two-component Gaussian Mixture Model was fit to the log1p-transformed UMI counts for each antigen (D1-4) (*80*). A cell was classified as positive if it belonged to the higher-mean component with a posterior probability ≥90% and its value exceeded the 99% percentile of the background distribution, which was derived from the three non-DENV (negative control) barcodes, for one (D1-4) or more E dimers (CR2-4) (Fig. S9A). Sequence similarity between CDR3 amino acid sequences identified by RATP-Ig and LIBRA-seq was assessed using blastp with an 1 × 10^−5^e-value threshold to identify putative matches.

### Statistical Analyses

To characterize antibody-binding profiles in an unsupervised manner and evaluate potential differences in antigen-binding preferences between single-stained and multiplex (cocktail)-stained assay formats, we conducted principal coordinates analysis (PCoA) and k-means (k=3) clustering using log10-transformed, geometric mean fluorescence intensity (gMFI) values. Pairwise dissimilarities between antibody profiles were calculated using Manhattan (L1) distance, defined as the sum of absolute differences across antigen-specific binding measurements. Larger Manhattan distances therefore indicate greater dissimilarity in antibody-binding profiles, whereas a distance of zero indicates identical values across all included antigens. Antigen-specific gMFI values were fitted post hoc to the PCoA ordination using envfit (R vegan package). For each antigen, the fitted vector indicates the direction of increasing gMFI values, and its length is proportional to the strength of the association between antigen-specific binding and the displayed PCoA configuration (first two PCoA axes). Antibody profiles that project farther in the direction of an antigen vector exhibit relatively higher binding to that antigen, whereas profiles projecting in the opposite direction exhibit relatively lower binding. Statistical significance of each fitted vector was assessed using permutation testing.

Elastic net regression was used for selection of BCR features (CDRH3 physicochemical properties and *IGHV* gene family) important for predicting FL-sensitivity of the sequences identified from DEN24 by RATP-Ig using the glmnet in the train function from the caret package in R (*81, 82*). Additionally, we used elastic net regression to identify important physicochemical properties for distinguishing the epitope targets of the CDRH3 sequences identified from DEN24 by RATP-Ig and 173 publicly available human α-DENV antibody sequences (*51*). FL-sensitive antibodies from DEN24 and publicly available α-FL antibody sequences were combined to represent “FL-like” antibodies during analysis. Model performance was evaluated by cross-validation using the glmnet framework in R, with hyperparameters tuned across values of alpha and lambda (selected model: alpha=0.90, lambda=0.02). The model achieved an overall accuracy of 58.2% (SD = 0.042), but this masked class imbalance in performance, consistent with the low Kappa score of 0.21 (SD = 0.09), which accounts for chance agreement across classes. Sensitivity for FL-like was high (94.7%) but with low specificity (25%), sensitivity for FL-agnostic antibodies was low (3.1%) but with high specificity (99%), and sensitivity for α-EDE was moderate (54.4%) with high specificity (94%). These results indicate the model is biased toward predicting the FL-like class and almost never predicts FL-agnostic antibodies. However, it rarely calls an antibody α-EDE if it is not a true α-EDE antibody and correctly identifies about half of true α-EDE antibodies.

We then combined these two approaches to classify sequenced DENV-specific cells (LIBRA-seq). For unsupervised characterization of the binding features of these cells and the identification of potential antigen-binding preferences, we performed PCoA with k-means clustering (k=5) on log1p-transformed, background-subtracted UMI counts. Manhattan distance was used to quantify dissimilarity among binding profiles. The trained elastic net classifier was used to assign the BCR sequences to predicted binding categories, including FL-like, FL-agnostic, and EDE-like antibodies. These assignments should be interpreted as prioritization hypotheses rather than definitive functional annotations and require experimental validation.

## Data Availability

The data and code generated for this study will deposited on Zenodo and made available at the time of publication.

## Acknowledgements

We would like to acknowledge David Ambrozak at the NIH Vaccine Research Center’s Flow Cytometry Core Facility for his assistance with FACS sorting, Liya Muslkinka from NIAID’s Research Technologies Branch Structural Biology core for her help with BLI, and NIAID’s Research Technologies Branch Flow Cytometry core for use of their facilities, including assistance from Melanie Cohen and Julie Laux. The 4E11 antibody was kindly shared with us by Sandra Mayer and Greg Gromowski (WRAIR). The cell lines and viruses used in this study were generously shared with us by the following: ZIKV, WNV, JEV, and ZIKV viral strains shared by Stephen Whitehead (NIH); DENV clinical isolates shared by Aravinda de Silva (University of North Carolina at Chapel Hill); Vero cells shared by Eva Harris (University of California, Berkeley); and the VeroFurin-Clone-1 cells shared by Ralph Baric (University of North Carolina at Chapel Hill).

## Funding

This research was supported by the Intramural Research Program of the National Institutes of Health (NIH); the Center for Human Immunology, Autoimmunity, and Inflammation (NIH); the NIH ReVAMPP grant U19AI181960-01, and NIH grant P01AI106695. The contributions of the NIH authors were made as part of their official duties as NIH federal employees, are in compliance with agency policy requirements, and are considered Works of the United States Government. The findings and conclusions described here are those of the authors and do not necessarily represent the views of the NIH or the U.S. Department of Health and Human Services.

## Author Contributions

Conceptualization: KEL, RAA, TSJ, DCD, LCK

Methodology: KEL, RAA, SS, DJT, SJ, LCK, RA, JJP, FC, ID

Investigation: KEL, RAA, SS, YB, SF, GB, DJT, PM, ARH, JST, JTR, LE, SJ, RA, JJP, FC, ID

Visualization: KEL, RAA, SJ, LCK

Funding acquisition: AMD, DCD, LCK, JC

Project administration: KEL, RAA, KG, LCK

Supervision: CAS, ED, BJD, JIC, ID, BAB, JLH, AMD, DCD, LCK

Writing – original draft: KEL, RAA, LCK Writing – review & editing: all authors

## Competing Interests

Authors declare that they have no competing interests.

## Data and Materials Availability

The data and code generated for this study will be deposited on Zenodo and made available at the time of publication.

## Notes

### Competing Interest Statement

The authors have declared no competing interest.

### Clinical Trial

NCT01306084

### Author Declarations

Ethical approval was obtained from the Institutional Review Board of the National Institutes of Health and is registered at clinicaltrials.gov (NCT01306084).

