## Supplementary Materials for "Signatures of the neutralizing dengue virus-reactive memory B cell repertoire identified by multiplexed envelope dimer probes"

#### Materials and Methods

**Virus Strains and Cell Lines.** Low-passage, representative dengue viruses 1-4 (DENV1-4) and Zika virus (ZIKV) viral isolates as well as vaccine strains for West Nile virus (WNV), Japanese encephalitis virus (JEV), and yellow fever virus (YFV) used in virus neutralization assays are listed as follows: DENV1/SriLanka/2014/GS0736, DENV2/SriLanka/2016/GS1816, DENV3/SriLanka/1989/UNC3001, and DENV4/SriLanka/1992/280056 (GenBank numbers: PQ667803, PQ667804, PQ667805, and PQ667806); ZIKV/Paraiba/2015 (GenBank number AF326573); WNV/DENV4Δ30 (WNV envelope [E] and pre-membrane [prM] in an attenuated DENV4 backbone) (*70, 71*); JEV SA14–14-2 (GenBank number MH258849); and YFV-17D (GenBank number X03700.1). Viruses were grown in wildtype Vero cells (CCL-81, ATCC) or furin-overexpressing Vero cells (VeroFurin-Clone-1) (*72*) to generate standard and mature viral stocks, respectively. Virus stocks were prepared by infecting Vero or VeroFurin-Clone-1 cells at a multiplicity of infection of 0.01 for one hour at 37ºC, removing the virus and gently washing the cells with sterile Phosphate Buffered Saline (PBS) without magnesium chloride or calcium chloride (Gibco), and culturing the cells in OptiPro™ Serum Free Medium supplemented with 4mM L-glutamine (Gibco) for 3-4 days at 37ºC and 5% $CO_2$ with 90% humidity. Viral supernatants were harvested at the first signs of cytopathic effects, stabilized with 1x Sucrose-Phosphate-Glutamate, flash frozen, and stored at -80ºC.

**Dengue virus (DENV)-Specific B Cell Sorting using a stabilized DENV2 E dimer protein with a partial knockdown of the FL (ΔFL D2) (Donor DEN20).** For our initial validation run with DEN20, isolated B cells were stained with 5μg/mL of ΔFL D2 E dimer, for 30 minutes before adding 5µg/mL of α-His-APC (J095G46, BioLegend) for 20 minutes. Cells were then incubated for another 30 minutes with 5µg/mL of DENV2 E-dimer before adding 5µg/mL of α-His-FITC (J095G46, BioLegend) and the B cell phenotyping antibody panel. The B cell phenotype was determined by staining with the following antibodies: CD19-PECy7 (J3-119, Beckman Coulter); CD20-BUV805 (2H7, BD Biosciences); IgD-BV785 (1A6-2, BioLegend); IgM-BUV395 (G20-127, BD Biosciences); IgG-PECF594 (G18-145, BD Biosciences). Dead cells, T cells, monocytes, and NK cells were excluded using the LIVE/DEAD Fixable Blue Dead Cell Stain (Molecular Probes, Invitrogen) and the antibodies: CD3-BV510 (OKT3, BioLegend), CD14-BV510 (M5E2, BioLegend), CD16-BV510 (3G8, BD Biosciences), and CD56-BV510 (HCD56, BioLegend). Gates for $IgG^{+}$ memory B cells specific for the E dimer ($APC^{+}FITC^{+}$) were set using cells from the DENV naïve donor.

**DENV E Dimer ELISA Screening.** RATP-Ig supernatants and purified monoclonal antibodies were screened for DENV-reactivity using a sandwich ELISA using biotinylated D1-4 (*40, 41*) and ΔFL D2 E dimers (*40*) to enable detection of quaternary binding antibodies as described previously (*83*). RATP-Ig supernatants were screened at a 1:10 dilution in technical duplicate and purified monoclonal antibodies were screened in technical triplicate at 10 µg/mL.

High binding microwell plates (Greiner Bio-One) were coated with 100ng of mouse α-His monoclonal antibody (OTI2B5, OriGene Technologies) in tris-buffered saline (TBS) overnight at 4ºC. Plates were washed with TBS + 0.05% Tween 20 (Sigma-Aldrich) (TBST) and blocking with TBST + 3% instant nonfat dry milk (Carnation) for 1h in a 37ºC shaking incubator. Plates were washed with TBST and incubated with 100ng of E dimer in TBST + 3% milk for 1h in a 37ºC shaking incubator. Again, plates were washed and 50µL of either RATP-Ig supernatants diluted 1:10 in TBST + 3% milk or 10µg/mL of purified monoclonal antibodies were added to wells and incubated for 2h in a 37ºC shaking incubator. Following incubation, plates were washed with TBST and then stained with 50µL of goat α-human IgG-AP antibody (Sigma-Aldrich) diluted 1:2000 in TBST for 1h in a 37ºC shaking incubator. Plates were washed and developed for 15min at room temperature with 50µL of pre-dissolved SIGMAFAST™ p-Nitrophenyl phosphate set (Sigma-Aldrich) before quenching with 50µL of 1N NaOH (Fisher Scientific). Absorbance at 405nm was measured using a Cytation7 (BioTek), and the relative OD (rel. OD) was calculated by subtracting the average background signal (blank) from all samples and then normalizing the average sample OD to the average OD for the α-HIV antibody VRC01 (*84*) by subtraction. E dimer binding breadth groupings were determined by a cutoff of a rel. OD greater than the mean +1 SD of the rel. OD of the blank (no cell) RATP-Ig supernatants for each E dimer, and the corresponding percentages for each group are plotted. Antibodies with binding to more than one E dimer serotype were classified as cross-reactive (CR), followed by the number of serotypes they bound to (CR2-4). FL-sensitivity (ΔFL D2 binding status) was determined using a ΔFL D2 dimer binding greater than the mean +1 SD Rel. OD of the blank (no cell) RATP-Ig supernatants.

**Standard and Mature Flavivirus Neutralization Assays.** Sera, RATP-Ig supernatants, and purified monoclonal antibodies were screened for neutralization of standard and mature flaviviruses by modifying a DENV Focus Reduction Neutralization Test (FRNT) as described previously (*6, 85, 86*). For initial RAPT-Ig supernatant screening, supernatants were screened using a 1:20 point dilution run in technical triplicate. For deeper characterization of the top neutralizing RATP-Ig supernatants, a 6 step, four-fold dilution series was used in the assay. Serum or plasma profiling was conducted using a 12 step, 2-fold dilution series starting at a final serum or plasma dilution of 1:10. Assays involving purified monoclonal antibodies were run in a 12 step, 3-fold dilution series starting at a final antibody concentration of 5 or 10 µg/mL.

Standard Vero-grown or mature Vero-Furin-Clone-1-grown viruses (60-80 foci per well) were titrated against antibody samples diluted in Opti-MEM (Gibco) supplemented with 2% FBS, 0.5% human albumin (NIH DVR Pharmacy), and 50µg/mL gentamicin (ThermoFisher Scientific) and incubated at 37°C for 60 minutes as described previously (*2-4*). After, 30µL of the sample-virus mixture was added in technical duplicate to 96-well flat bottom plates (Corning) seeded with Vero cells 24h prior, incubated for 1h at 37°C, and then 150µL of warmed 1% methylcellulose in supplemented Opti-MEM overlay medium was added to cells. Plates were incubated for 40h (standard JEV and ZIKV), 48h (mature DENV2 and standard/mature DENV4), 50h (standard WNV and YFV), or 52h (standard/mature DENV1, standard DENV2, standard/mature DENV3) at 37°C before overlay was washed off and cells were fixed with ice-cold 80% methanol and stored at -80°C. Plates were thawed and immunostained with 100µL of 0.4µg/mL mouse α-flavivirus antibodies 2H2 and 4G2 (except 1:3000 of JEV immune ascitic fluid (VR-1259AF, ATCC) was used for JEV assays) for 2h at 37°C. Plates were counterstained with 100µL of peroxidase-labeled goat anti-mouse IgG (SeraCare) diluted 1:3000 in PBS+5% milk for 1h at 37°C and developed with 30µL of TrueBlue Peroxidase (SeraCare) substrate until foci were clearly distinguishable (5-20 minutes). Plates were imaged using an ImmunoSpot Analyzer (Cellular Technology Limited), and foci were counted using the Viridot package in R (*2*).

Both $FRNT_{50}$ titers and antibody $EC_{50}$ values were estimated by a 50% reduction in foci relative to virus only wells using a 4-parameter log-logistic regression with constraints (upper=100 and lower=0) using the drc package in R (*87*). DENV $EC_{50}$ values above the limit of detection were estimated from the logistic regression curves for concentrations between 5,000-10,000 ng/mL, and values exceeding the 10,00 ng/mL were set as a two-fold concentration above the limit of detection. For $FRNT_{50}$ titers below the limit of detection, values were set as a two-fold dilution below the limit. As controls, each strain was titrated against known α-DENV monoclonal antibodies (e.g. 1M7 (*28*), EDE-C8 (*15*), EDE-C10 (*15*), and EDE-B7 (*15*)), predetermined neutralizing sera and/or plasma against a flavivirus panel, as well as the α-HIV antibody, VRC01 (*84*), to control for non-specific neutralization effects.

**Biolayer Interferometry (BLI).** BLI experiments were carried out using the Octet R8 system with D2 dimer immobilized on Ni-NTA biosensors (Sartorius, #18-5101) and antibodies as analytes. Antibodies were prepared by two-fold serial dilutions in assay buffer (0.1% BSA, 0.2% Tween-20, 1X PBS pH 7.4) ranging from 50-6.25 nM for our top neutralizing FL-sensitive (excluding DEN24-1G02) and FL-agnostic antibodies, the α-EDE antibodies (C8, C10, and B7 (*15*)), and the α-FL antibody 1M7 (*28*). BLI experiments were performed at 37°C with agitation at 1,000 rpm, and the four concentrations of each antibody were run in duplicate. D2 dimer at a concentration of 1.25 μg/mL was loaded on the sensors for 300 s. After dipping into assay buffer for 120 s (baseline), biosensors were dipped in the 2-fold dilutions of various antibodies (association) for 300 s, followed by dipping into assay buffer for 300 s (dissociation). Data analysis and curve fitting of the four concentrations of each antibody were performed using Octet Analysis Studio 13 software to obtain the binding kinetic constants.

**Epitope Mapping.** Epitope mapping was performed as described previously (*44*). Briefly, alanine scanning mutagenesis libraries for DENV2/Thailand/16681/84 (*44*) and ZIKV/Brazil/SPH2015 (*88*) prM/E were expressed individually with or without furin protease in HEK-293T cells for 22h. Afterwards, cells were intracellularly stained with purified monoclonal antibodies (0.1-2.0 μg/mL) and then counterstained with 3.75 μg/mL Alexa Fluor 488-conjugated secondary antibody (Jackson ImmunoResearch Laboratories). Cells were washed and mean cellular fluorescence was detected using an Intellicyte iQue flow cytometer (Sartorius). Antibody reactivity against each mutant clone was calculated relative to signal observed for wild-type prM/E by subtracting the signal from mock-transfected samples and normalizing to the signal from wild-type-transfected controls. Mutations within clones identified critical residues for the monoclonal antibodies' epitope when the test monoclonal antibody was unable to recognize that mutant (<20% binding), but other control antibodies were still able to bind. Secondary clones do not pass the <20% binding threshold but still demonstrate binding activity that is sufficiently reduced (<27%) to suggest that they contribute to antibody binding.

**Antigen-Specific B Cell Immunophenotyping.** Antigen probes were conjugated to their respective fluorophores as described above. PBMCs were thawed, washed with twice with PBS before staining each sample with 22.5 µL of a cocktail containing 5 µL/sample of Human TruStain FcX™ (BioLegend) and Zombie NIR™ Fixable Viability Kit (BioLegend) at a final staining dilution of 1:1600 in PBS for 15 min at room temperature, protected from light. Without washing, 17.5 µL of a cocktail containing chemokine receptor antibodies (α-CXCR3, CXCR4, CXCR5, and CCR7) at the dilution indicated in Table S2 in 10 µL/sample of BD Horizon™ Brilliant Stain Buffer Plus (BD Biosciences) were added per sample and incubated for 15 min at a 37°C protected from light. Next, without washing, the samples were stained with 10 µL of pooled D1-4 dimer cocktail in PBS+2% FBS were added for 30 min at 4°C protected from light. Finally, 56 µL of a cocktail containing the remaining surface markers (Table S4) diluted in 10 µL/test of BD Horizon™ Brilliant Stain Buffer Plus, 5 µL/test of True-Stain Monocyte Blocker™ (BioLegend), 5 µL/test of CellBlox™ Plus Blocking Buffer (Invitrogen), and PBS+2% FSB were added to unwashed cells and stained for another 30 min at 4°C protected from light. Following staining, the cells were washed twice with PBS+2% FBS and then fixed for 15 min with 4% paraformaldehyde in PBS at 4°C. Fixation buffer was replaced with PBS+2% FBS, and cells were stored at 4°C overnight. Samples were acquired next day on a Sony ID7000 spectral analyzer. Single color controls used for spectral unmixing were stained the morning of the acquisition using UltraComp eBeads™ Plus beads (Invitrogen). Controls for antigen probes were made by labeling beads with a biotinylated α-CD16 antibody (3G8, BioLegend) before staining with the fluorescently labeled streptavidin tags used to label each antigen.

**Flow Cytometric Analyses.** Exported FCS files were pre-processed using the PeacoQC (*91*) plugin in FlowJo 10.10.0 (BD Biosciences). FCS files for passing events were imported into the OMIQ software from Dotmatics (www.omiq.ai,www.dotmatics.com) for all remaining analyses. Data were scaled and manually gated to remove debris, protein aggregates, doublets, dead cells, and CD45 negative events. UMAP analysis was used to visualize $CD19^+$ cell populations for DEN12 (*92, 93*). UMAP settings were as follows: all fluorescent parameters except CD45-SBUV510, Zombie NIR, and the antigen probes (D1, D2, D3, D4, and CoV2); Neighbors = 20; Minimum Distance = 0.6; Components = 2; Metric = Euclidean; Learning Rate = 1; Epochs = 200; Random Seeds = 5488; and Embedding Initialization = spectral. FlowSOM was then run to cluster the data (*94*), with settings as follows: clustering features included all fluorescent parameters besides CD45, Zombie NIR, and the antigen probes; umap_1 and umap_2; xdim = 12 and ydim = 12; rlen = 10; Distance Metric = Euclidian; consensus metaclustering with k = 12; Random Seed = 9639. Metaclusters were annotated by marker expression profiles. DENV-positive cells were identified by manual gating against samples stained with D1-4 dimer minus one of the dimers and mock HSA cocktail staining controls, and Boolean combination gating was applied to define E dimer reactivities.

#### Figs. S1 to S10

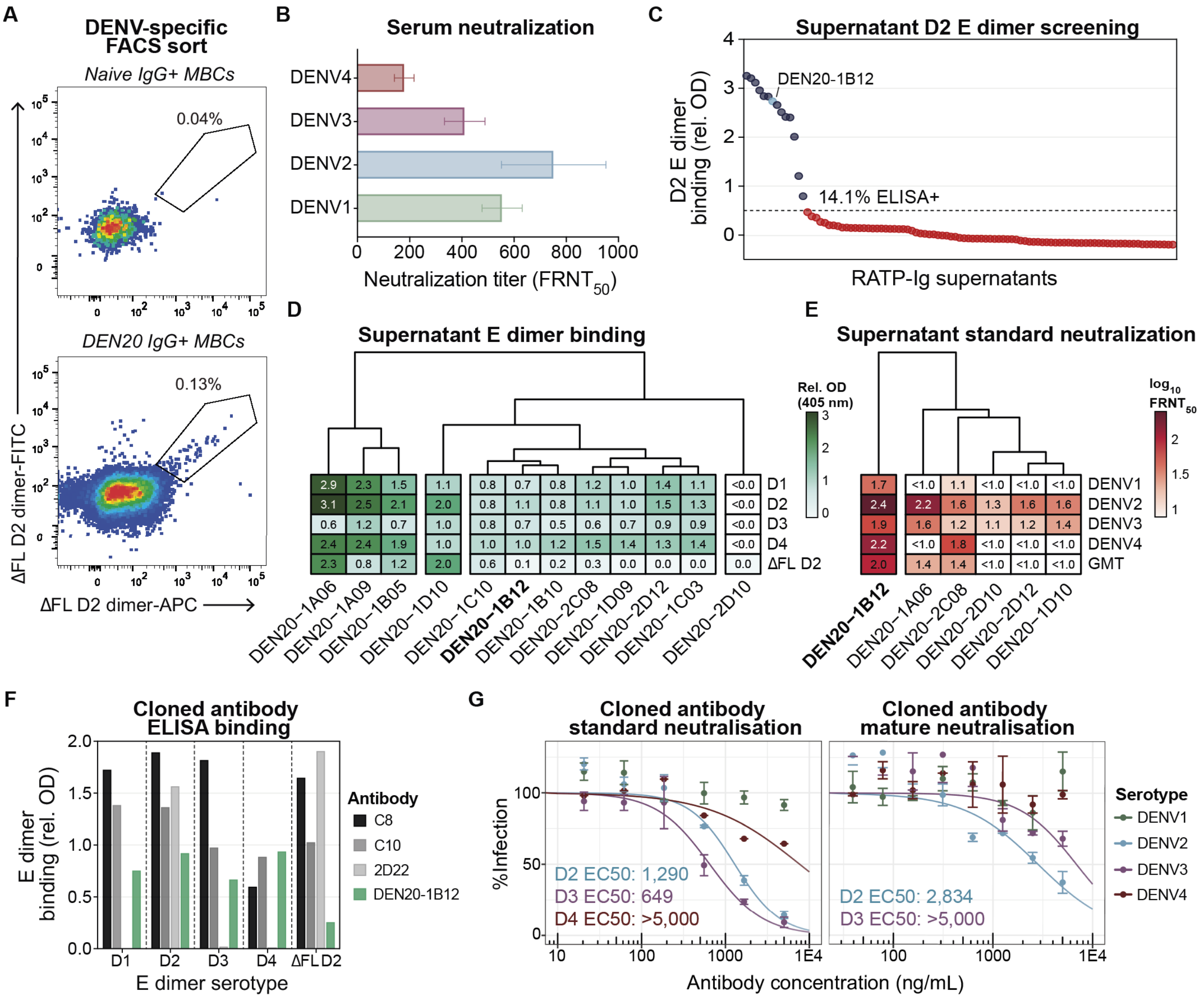


**Fig. S1: ΔFL D2 dimer probe captures memory B cells encoding DENV-binding and -neutralizing B cell receptors (BCRs) from an individual six weeks after infection. (A)** ΔFL D2 dimer-specific IgG+ memory B cell frequencies for DENV-immune donor DEN20 used in a RATP-Ig pilot, compared to a DENV-naïve donor (sort gate control). **(B)** Standard DENV1-4 focus reduction neutralization ($FRNT_{50}$) titers for DEN20 serum at enrollment determined by FRNT. Bar graph shows median $FRNT_{50}$ with interquartile range error bars. **(C)** D2 dimer ELISA screen for DEN20 RATP-Ig supernatants with recovered BCR heavy and light chain sequences.

**(D)** Heatmap of relative OD for D1-4 and ΔFL D2 dimers from DEN20 RATP-Ig supernatants with OD >2 in C. **(E)** Heatmap of standard DENV1-4 $log_{10}FRNT_{50}$ titers for representative DEN20 RATP-Ig supernatants from the four E dimer binding clusters in D. **(F)** Relative D1-4 and ΔFL D2 dimer binding for 10μg/mL purified DEN20-1B12 versus known DENV2 type-specific (2D22) and broadly neutralizing EDE1 (C8, C10) antibodies. **(G)** Standard (left) and mature (right) DENV neutralization curves for purified DEN20-1B12. **Abbreviations** – MBCs: memory B cells; Rel. OD: relative optical density; D1-4: DENV1-4 E dimer; GMT: DENV1-4 geometric mean titer; EC50: half-maximal effective neutralization concentration.

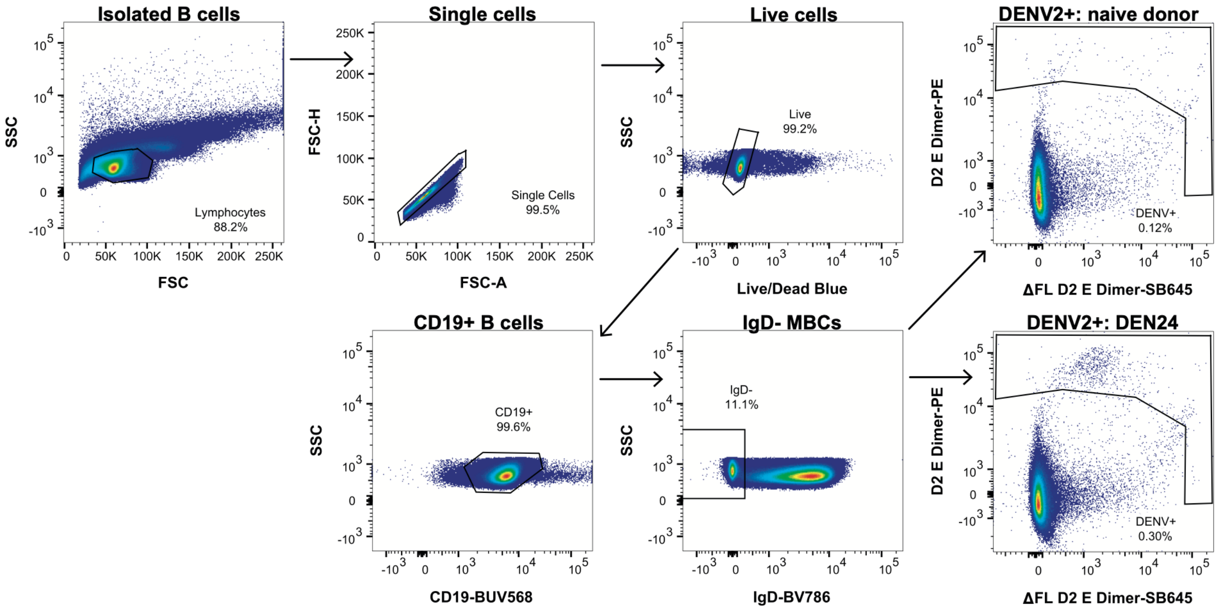


**Fig. S2: Gating scheme of the DENV-specific, single cell sort for DEN24 using the D2 and ΔFL D2 E dimer probes.** B cells were isolated from PBMCs, stained with D2 dimer-PE and ΔFL D2 dimer-SB645, and DENV-specific IgD$^{-}$ memory B cells were sorted positive. The DENV2$^{+}$ sort gate was set using a DENV-naïve individual as a control.

A

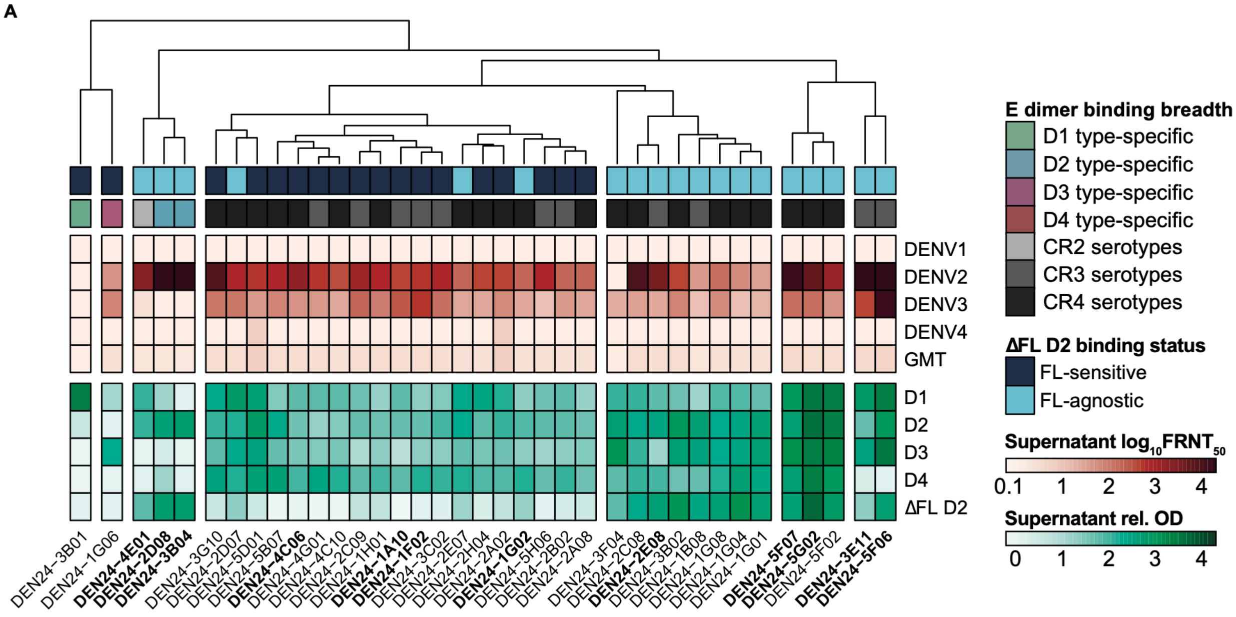


B

| Antibody | Heavy Chain Gene Use | | Light Chain Gene Use | | %SHM | | CDR3 Length | | CDRH3 Amino Acid Properties | | | | E Dimer Binding | | | | |
|---|---|---|---|---|---|---|---|---|---|---|---|---|---|---|---|---|---|
| | V | J | V | J | $V_H$ | $V_L$ | HC | LC | Polarity | Charge | %Acidic | %Aromatic | D1 | D2 | D3 | D4 | ΔD2 |
| DEN24-4E01 | IGHV1-3*01 | IGHJ5*02 | IGLV7-43*01 | IGLJ3*02 | 10.4 | 3.3 | 15 | 11 | 8.85 | 0.00 | 13.3 | 13.3 | | | | | |
| DEN24-5F07 | IGHV3-30-5*01 | IGHJ6*02 | IGKV3-11*01 | IGKJ1*01 | 10.8 | 4.9 | 10 | 11 | 8.74 | 0.00 | 10.0 | 0.0 | | | | | |
| DEN24-5F06 | IGHV4-31*03 | IGHJ6*02 | IGKV3D-15*01 | IGKJ3*01 | 11.7 | 5.3 | 23 | 13 | 8.79 | -0.01 | 13.0 | 17.4 | | | | | |
| DEN24-5G02 | IGHV4-61*02 | IGHJ1*01 | IGKV3-15*01 | IGKJ1*01 | 8.9 | 5.3 | 16 | 12 | 8.19 | 0.85 | 6.3 | 18.8 | | | | | |
| DEN24-2D08 | IGHV1-18*01 | IGHJ6*03 | IGLV2-11*01 | IGLJ3*02 | 8.3 | 4.8 | 20 | 11 | 8.07 | -1.01 | 10.0 | 30.0 | | | | | |
| DEN24-3B04 | IGHV1-69*01 | IGHJ4*02 | IGKV3-11*01 | IGKJ2*01 | 10.1 | 4.9 | 15 | 11 | 9.36 | 0.22 | 13.3 | 20.0 | | | | | |
| DEN24-1G02 | IGHV3-33*05 | IGHJ4*02 | IGLV1-51*01 | IGLJ3*02 | 11.5 | 6.4 | 14 | 13 | 8.70 | -2.00 | 21.4 | 28.6 | | | | | |
| DEN24-3E11 | IGHV4-39*01 | IGHJ4*02 | IGLV7-43*01 | IGLJ3*01 | 11.6 | 3.7 | 16 | 11 | 8.81 | -2.00 | 12.5 | 18.8 | | | | | |
| DEN24-2E08 | IGHV4-39*01 | IGHJ5*02 | IGKV1-9*01 | IGKJ3*01 | 10.7 | 4.9 | 10 | 11 | 8.92 | 0.00 | 10.0 | 30.0 | | | | | |
| DEN24-1F02 | IGHV4-39*02 | IGHJ4*02 | IGLV2-14*01 | IGLJ3*01 | 11 | 4.4 | 11 | 12 | 7.92 | 0.00 | 9.1 | 36.4 | | | | | |
| DEN24-1A10 | IGHV5-51*01 | IGHJ4*02 | IGKV3-15*01 | IGKJ3*01 | 7.3 | 1.1 | 17 | 10 | 9.15 | -1.89 | 17.6 | 23.5 | | | | | |
| DEN24-4C06 | IGHV5-51*01 | IGHJ5*02 | IGKV3-15*01 | IGKJ1*01 | 8.3 | 6.4 | 17 | 10 | 8.95 | -1.89 | 17.6 | 17.6 | | | | | |

Rel. OD
0 1.25 2.5

C

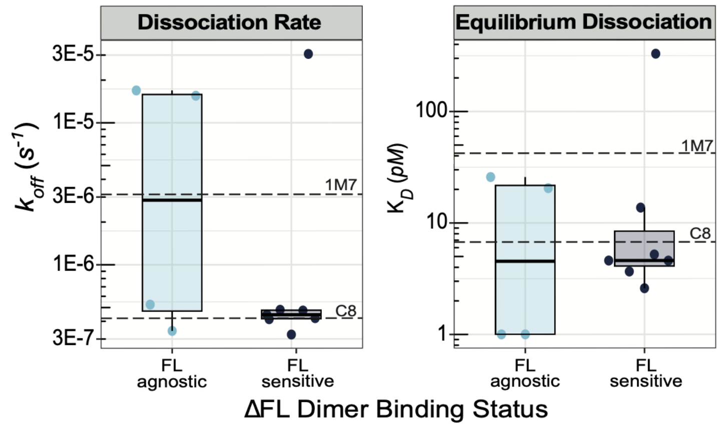


**Fig. S3. Screening and selection of top neutralizing antibodies. (A)** Summary heatmap of the average $\log_{10}$ $FRNT_{50}$ (top red section) of each antibody supernatant by mature FRNT and the average relative OD for D1-4 and ΔFL D2 dimer binding by ELISA (bottom green section). The annotations are based on the binding profile definitions from Fig. 1D&F. Antibodies selected for further exploration are indicated in bold. **(B)** Table indicating the VJ genes, somatic hypermutation rates, heavy (HC) and light (LC) chain physicochemical properties, and ELISA binding of the 12 neutralizing antibodies chosen for further exploration. Heatmap color gradation corresponds to E dimer binding intensity (Rel. OD at 10μg/mL) against D1-4 and ΔFL D2 dimers. **(C)** The $k_{off}$ rates ($s^{-1}$) and $K_D$ (pM) values for FL-agnostic and FL-sensitive antibodies against the D2 dimer determined by biolayer interferometry. The $k_{off}$ and $K_D$ values for 1M7 and C8 are indicated by dashed lines. No significant differences between FL-agnostic and FL-sensitive antibodies were detected using a Wilcoxon rank sum test. **Abbreviations** – CR2-4: Cross reactive to 2-4 serotypes;

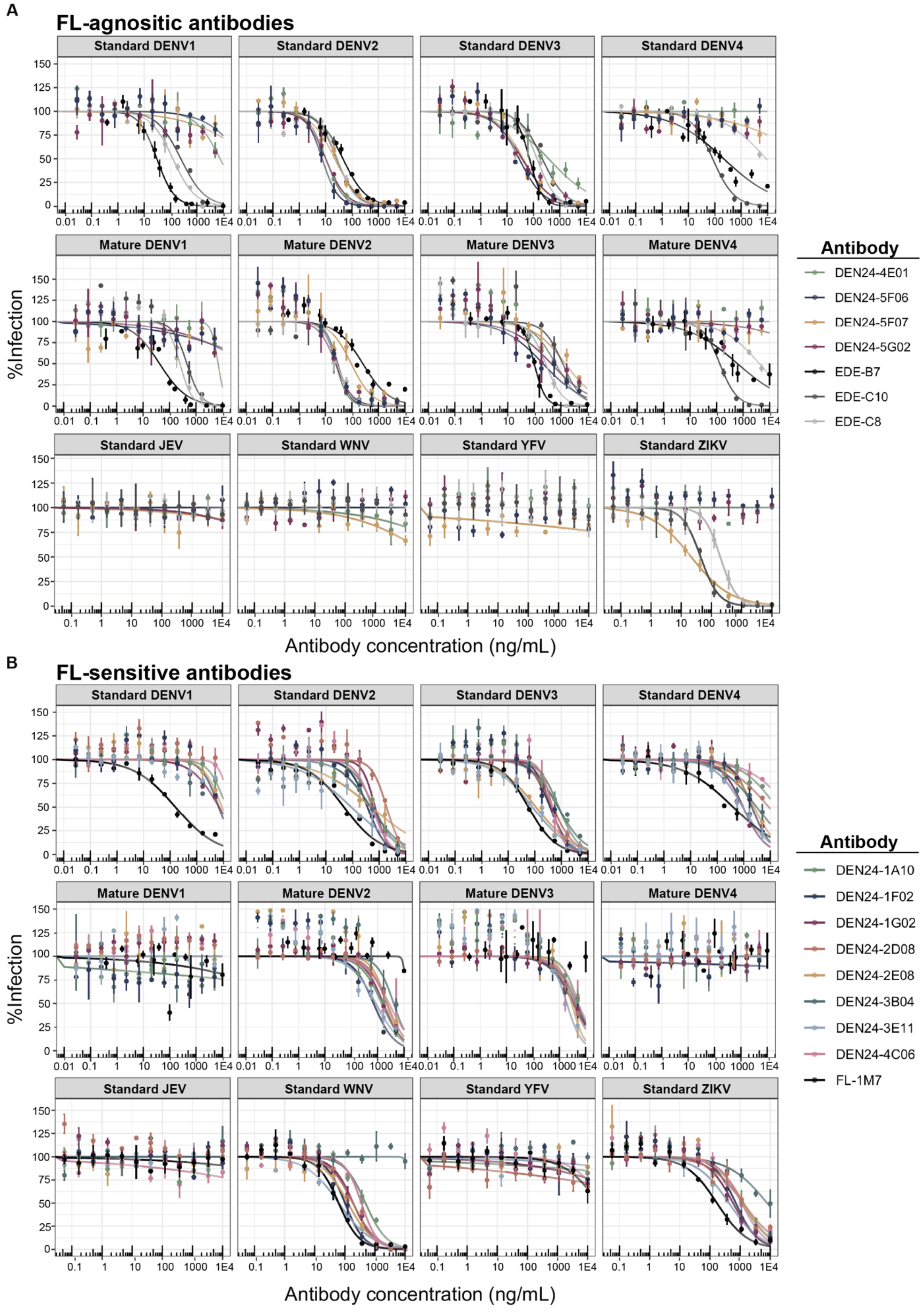
A
FL-agnositic antibodies
Standard DENV1
Standard DENV2
Standard DENV3
Standard DENV4
Mature DENV1
Mature DENV2
Mature DENV3
Mature DENV4
Standard JEV
Standard WNV
Standard YFV
Standard ZIKV
%Infection
Antibody concentration (ng/mL)
Antibody
DEN24-4E01
DEN24-5F06
DEN24-5F07
DEN24-5G02
EDE-B7
EDE-C10
EDE-C8
B
FL-sensitive antibodies
Standard DENV1
Standard DENV2
Standard DENV3
Standard DENV4
Mature DENV1
Mature DENV2
Mature DENV3
Mature DENV4
Standard JEV
Standard WNV
Standard YFV
Standard ZIKV
%Infection
Antibody concentration (ng/mL)
Antibody
DEN24-1A10
DEN24-1F02
DEN24-1G02
DEN24-2D08
DEN24-2E08
DEN24-3B04
DEN24-3E11
DEN24-4C06
FL-1M7

V: variable region; J: joining region.

**Fig. S4. Flavivirus neutralization curves for purified FL-agnostic and FL-sensitive antibodies.** Standard and mature DENV1-4 and non-DENV flavivirus (JEV, WNV, YFV, ZIKV) dose-response neutralization curves for **(A)** FL-agnostic and **(B)** FL-sensitive antibodies measured by FRNT on Vero cells. The known human antibodies EDE-B7, EDE-C8, EDE-C10, and FL-1M7 were included for reference.

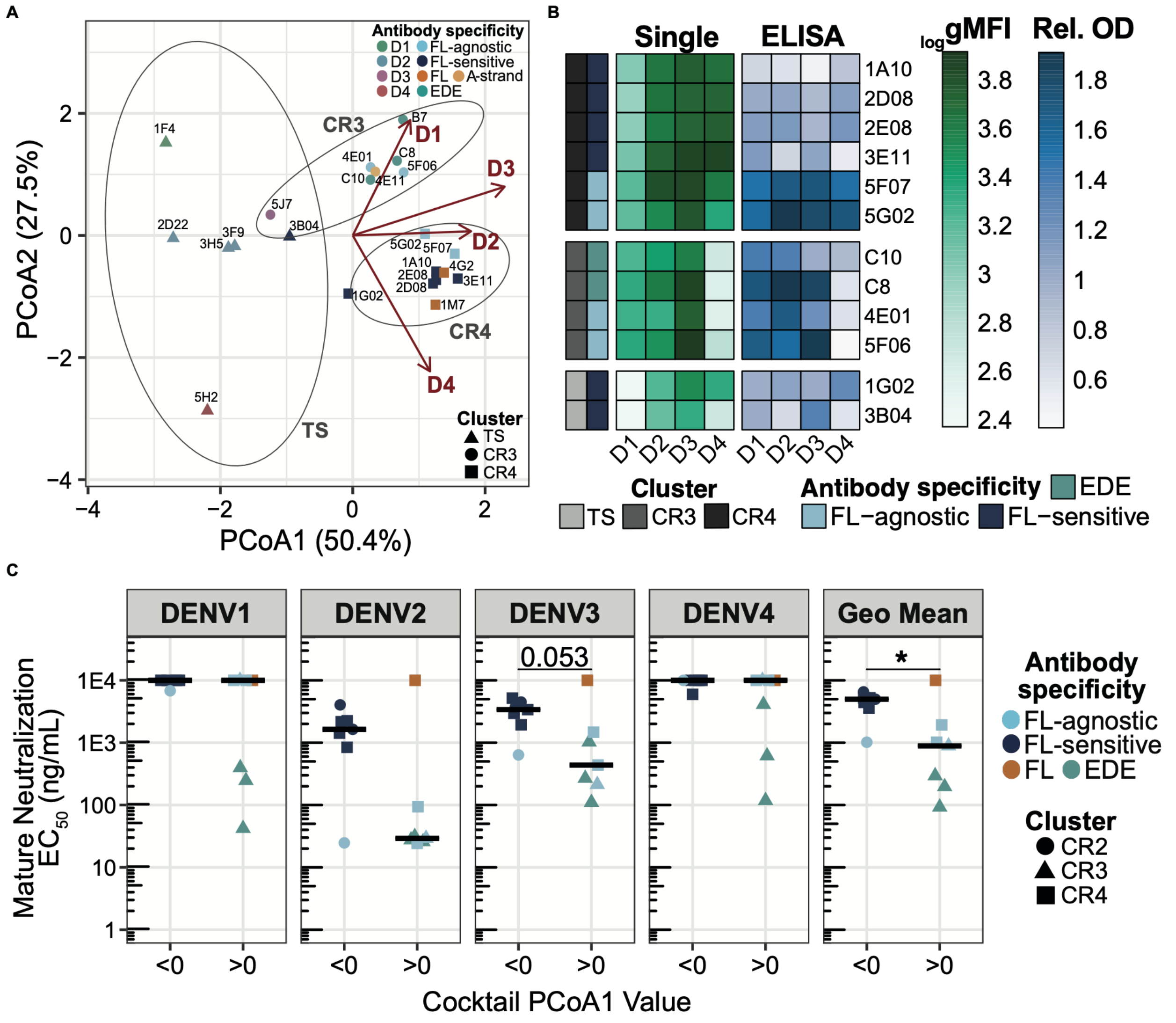

**Fig. S5. Single-stained cytometric bead binding analyses, and mature neutralizing antibody titers grouped by multiplexed bead assay binding profile. (A)** Principal coordinates analysis (PCoA) performed using Manhattan distances calculated from standardized single-stained D1-4 geometric mean fluorescent intensity (gMFI) values. Antibodies are shaped by their k-means cluster assignment (TS, CR3, and CR4) and colored by antibody specificity. Fitted D1-4 vectors indicate the direction of increasing gMFI values. Antibodies positioned closer to, and in the direction of, a given probe vector exhibit relatively higher gMFI values to that probe. **(B)** D1-4 dimer gMFI values from the single-stained cytometric bead assay and ELISA relative optical density (Rel. OD) values for cross-reactive and serotype-specific antibodies annotated by their antibody specificity and cluster identity from A. **(C)** Mature DENV neutralization $EC_{50}$ values for antibodies with PCoA1 values <1 or >1 in Fig. 4F. Crossbars represent median values. Points are colored by their antibody specificity and their shape denotes their cluster identity from Fig. 4F. Significance was evaluated by Wilcoxon rank-sum test.

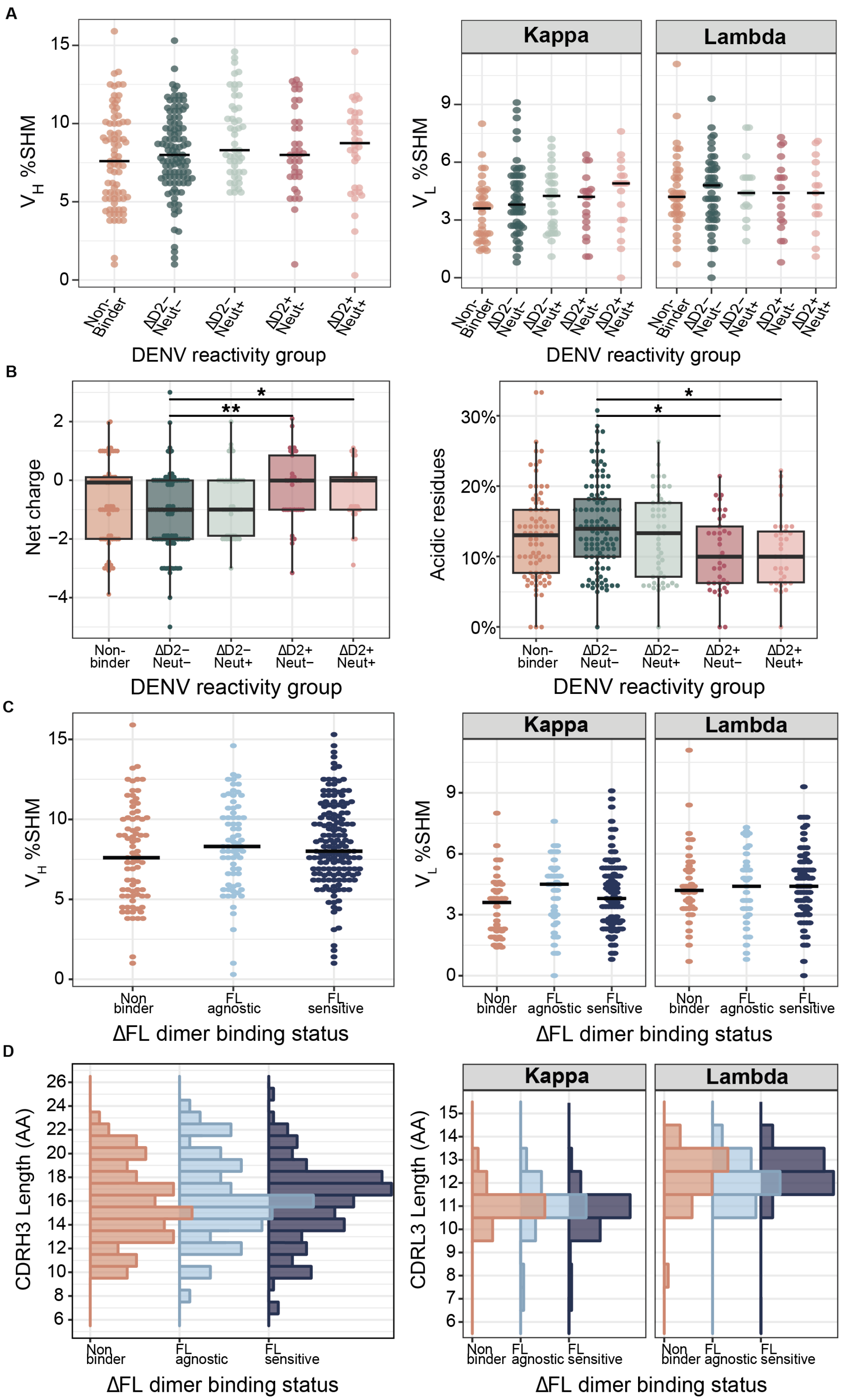
A
Kappa
Lambda
V_H %SHM
V_L %SHM
Non-Binder
ΔD2– Neut–
ΔD2– Neut+
ΔD2+ Neut–
ΔD2+ Neut+
DENV reactivity group
B
Net charge
Acidic residues
Non-binder
DENV reactivity group
C
V_H %SHM
V_L %SHM
Non binder
FL agnostic
FL sensitive
ΔFL dimer binding status
D
CDRH3 Length (AA)
CDRL3 Length (AA)
ΔFL dimer binding status

**Fig. S6. Somatic hypermutation rates and CDR3 lengths by DENV reactivity and ΔFL dimer binding. (A)** Somatic hypermutation rates (%SHM) for the heavy ($V_H$) and light ($V_L$) variable regions stratified by DENV reactivity group. **(B)** Net charge and acidic residue content of the heavy chain complementarity determining region 3 (CDRH3) for supernatants stratified by DENV reactivity. **(C)** $V_H$ and $V_L$ chain %SHM stratified by ΔFL dimer binding status. **(D)** $V_H$ and $V_L$ chain CDR3 lengths (number of amino acid [AA] residues) stratified by ΔFL dimer binding status. Significance was evaluated by Kruskal-Wallis test with post-hoc Dunn's test (Bonferroni-corrected). Median values for each group are represented as crossbars. *$p < 0.05$ and **$p < 0.01$, else no significance was observed.

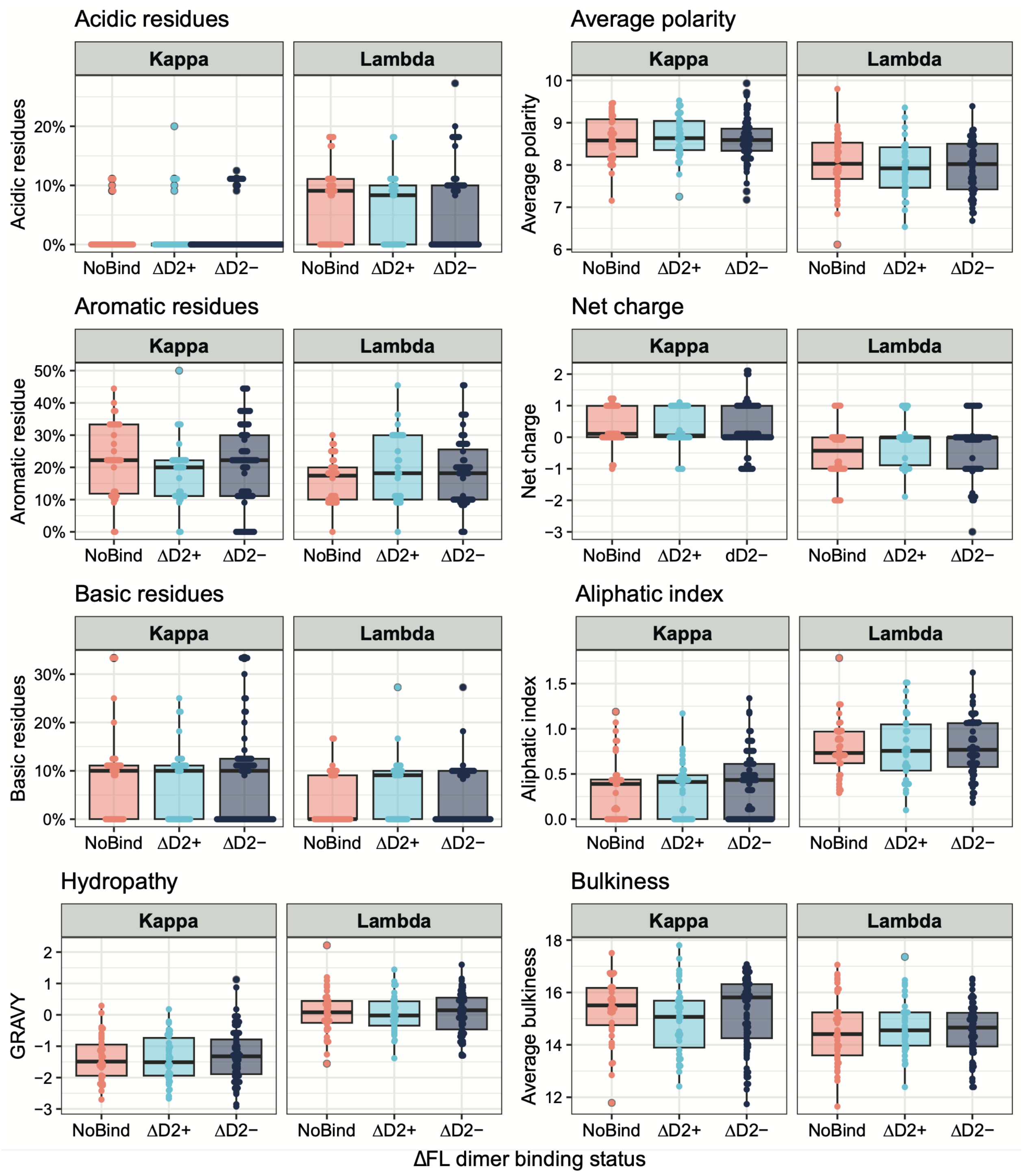


**Fig. S7. Physiochemical features of light chain complementarity determining region 3 (CDRL3) sequences by E dimer binding groups.** Physicochemical properties of the CDRL3 of isolated E dimer non-binding (NoBind), FL-sensitive (ΔD2-), or FL-agnostic (ΔD2+), antibodies.

Statistical significance was assessed using a Kruskal-Wallis test, and no significant differences between groups were observed. **Abbreviations** – GRAVY: grand average of hydropathy.

**A**

| E Dimer Construct | Fusion Loop AA Sequence | Hydropathicity (GRAVY) | Average Bulkiness | Average Polarity | Net Charge | Aliphatic Index | Basic Residues | Acidic Residues | Aromatic Residues |
|---|---|---|---|---|---|---|---|---|---|
| WT D2 | 98-DRGWGNGCGLFGK-110 | -0.71 | 11.37 | 8.65 | 0.93 | 3.0 | 15.38% | 7.69% | 15.38% |
| ΔFL D2 | 98-DRGWGNGCDLFGK-110 | -0.95 | 12.01 | 8.95 | -0.07 | 3.0 | 15.38% | 15.38% | 15.38% |

Small + Hydrophobic Polar + G Acidic Basic

**B**

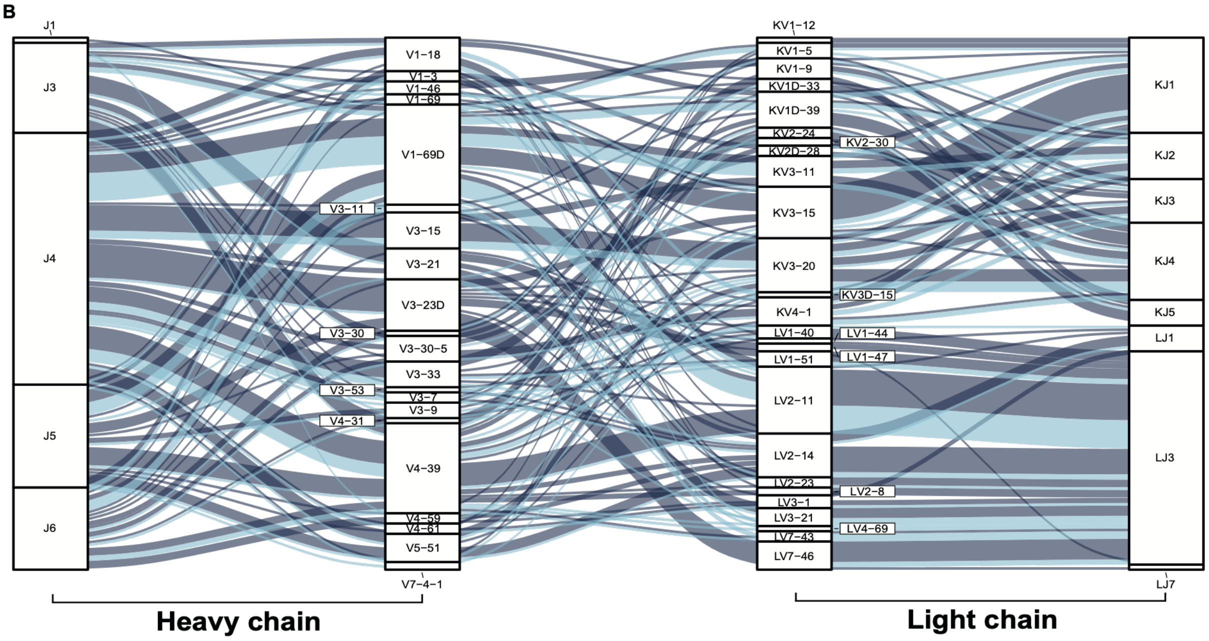


**Fig. S8. Fusion loop physicochemical properties and VJ pairings of DENV-binding cells. (A)** Comparison of the physicochemical properties of the wildtype (WT) D2 and ΔFL D2 E dimer fusion loops. **(B)** Paired heavy and light chain V-J combinations of E dimer binding clones represented as an alluvial plot. Strata height corresponds to the relative frequency at which V and J genes were identified, and ribbon width corresponds to the frequency of each V-J combination. Non-expanded clonotypes (size = 1) were not visualized. Ribbon color denotes ΔFL D2 dimer binding status of the clones for each pairing (navy blue = FL-sensitive and light blue = FL-agnostic).

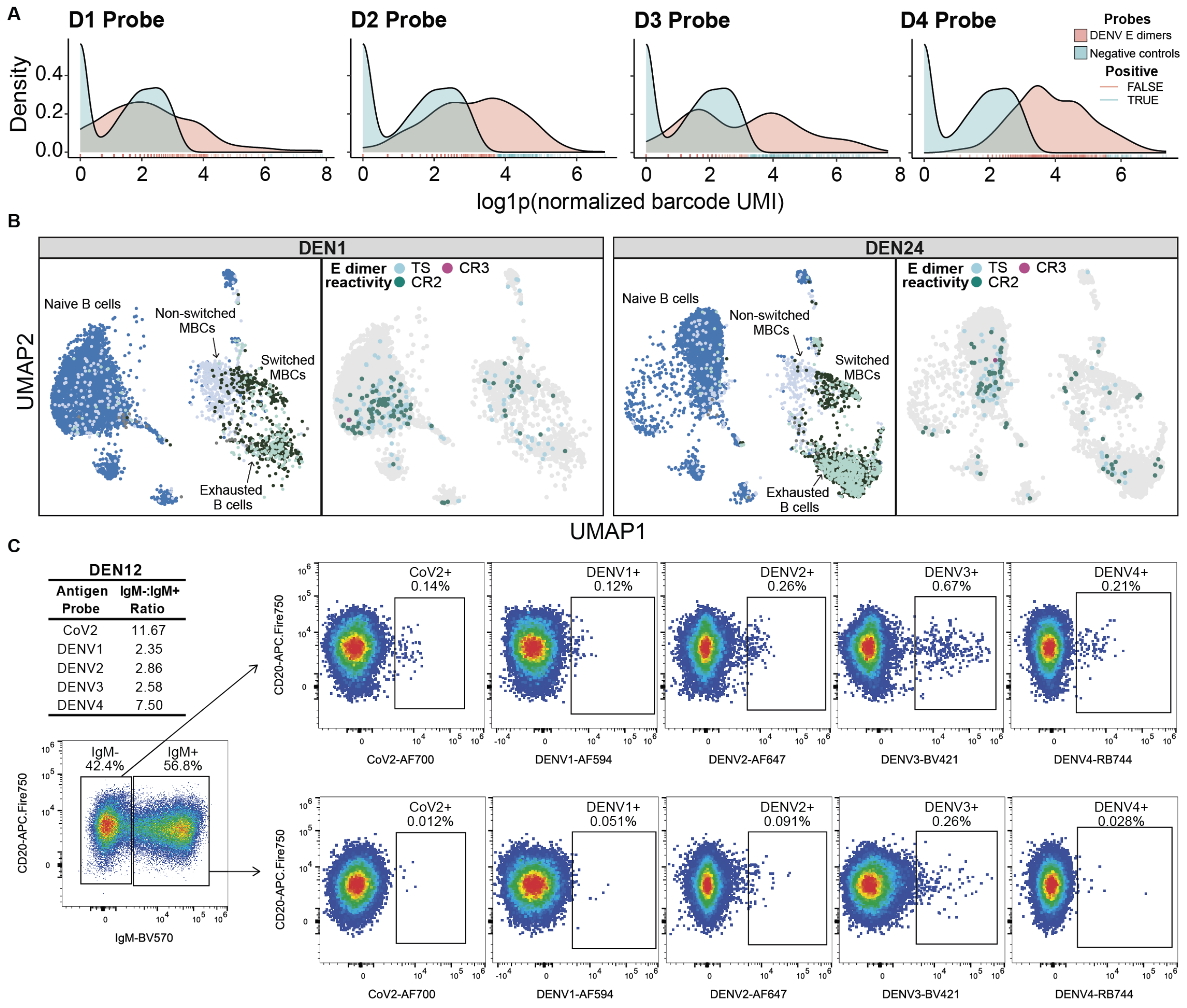


| DEN12 | |
|---|---|
| Antigen Probe | IgM-:IgM+ Ratio |
| CoV2 | 11.67 |
| DENV1 | 2.35 |
| DENV2 | 2.86 |
| DENV3 | 2.58 |
| DENV4 | 7.50 |

**Fig. S9**. **Multiplexed DENV1-4 probe staining profiles across assays. (A)** Density distribution of log-transformed UMI counts for each probe estimated using Gaussian Mixture Model to distinguish E dimer binding signal from background. The solid curve represents the model-estimated probability density, while tick marks along the x-axis indicate the distribution of individual data points. **(B)** UMAPs of B cells from DEN1 and DEN24 identified by LIBRA-seq in which each dot represents a cell, colored by cell subset annotation (left) and their E dimer reactivity (right). **(C)** DENV1-4 E dimer and SARS-CoV-2 D614G Spike (CoV2) gates for IgM⁻

(top) vs $IgM^+$ (bottom) B cells ($CD19^+CD20^+$) from DEN12 measured by flow cytometry. Table insert lists the $IgM^–$:$IgM^+$ staining ratio for each probe. **Abbreviations** – TS: serotype-specific; D1-4: D1-4 E dimer type-specific; CR2: cross-reactive to 2 serotypes; CR3: cross-reactive to 3 serotypes, CR4: cross-reactive to 4 serotypes; UMI: Unique molecular identifier.

### Tables S1 to S4

**Table S1:** LIBRA-seq clones sharing >80 % CDRH3 amino acid homology with RATP-Ig-identified antibodies from DEN24 and their corresponding RATP-Ig E dimer reactivity for homologous clones.[a]

| Donor ID | Heavy chain genes | | | CDRH3 alignment | Percent similarity | RATP-Ig HC match | Light chain genes | | CDRL3 alignment | Percent similarity | RATP-Ig LC match | Probe UMI counts | | | | | | Antigen reactivity | |
|---|---|---|---|---|---|---|---|---|---|---|---|---|---|---|---|---|---|---|---|
| | V | D | J | | | | V | J | | | | D1 | D2 | D3 | D4 | CoV2 | HSA | LIBRA-seq | RATP-Ig |
| DEN24 | V1-69*06 | D5-18*01 | J3*02 | AKVMNPSMGYNAFET<br>AKVMNPSMGYNAFET<br>*************** | 100 | DEN24-1G08 | KV1-39*01 | KJ2*01 | QQSYNSPPT<br>QQSYNSPPT<br>********* | 100 | DEN24-1G08 | 14 | **46** | **75** | 84 | 14 | 16 | CR2 | CR4 |
| DEN24 | V3-53*01 | D1-1*01 | J4*02 | ARVRDCSHINCLKIFDC<br>ARVRDCSHINCLKIFDC<br>***************** | 100 | DEN24-3H06 | KV1-5*05 | KJ4*01 | KQHAVHPLS<br>KQHAVHPLS<br>********* | 100 | DEN24-3H06 | 2 | 3 | 1 | 9 | 3 | 1 | Non-binder | Non-binder |
| DEN24 | V1-46*01 | D4/OR15-4a*01 | J5*02 | ARGGETAMVPTPVGAWFDP<br>ARGGETAMVPTPMGAWFDP<br>************:****** | 94.7 | DEN24-3C12 | KV1-9*01 | KJ5*01 | QQLNNYPVT<br>QQLNNYPVT<br>********* | 100 | DEN24-2H04 | 54 | 15 | **85** | 119 | 10 | 13 | D3 TS | HC: D4 TS<br>LC: CR4 |
| DEN12 | V4-39*01 | D3-10*03 | J5*01 | ARQRENWFDS<br>ARQLENWFDS<br>*** ****** | 90.0 | DEN24-3A05 | KV1-9*01 | KJ4*01 | QQLNNYVFT<br>QQLNNYVFT<br>********* | 100 | DEN24-2D10 | 39 | **54** | **31** | 88 | 13 | 18 | CR2 | HC: CR2<br>LC: CR3 |
| DEN24 | V1-69*01 | D3-10*01 | J4*02 | ARMKYGAGTYYNGRGHFDA<br>ARIKYGAGTYYNGRGHFDS<br>**:***************: | 89.5 | DEN24-4H09 | LV2-23*02 | LJ3*02 | CSYVGSSTWV<br>CSYVGSYTWV<br>****** *** | 90.0 | DEN24-1B08 | 23 | 20 | 22 | 70 | 3 | 13 | Non-binder | HC: CR4<br>LC: CR3 |
| DEN24 | V3-53*01 | D4-23*01 | J4*02 | ARVRDCGNVNCLKIFDC<br>ARVRDCSHINCLKIFDC<br>****** .:.******** | 82.4 | DEN24-3H06 | KV1-5*04 | KJ4*01 | KQHSLHPLS<br>KQHAVHPLS<br>***:.:**** | 77.8 | DEN24-3H06 | 1 | 1 | 0 | 8 | 8 | 4 | Non-binder | Non-binder |

[a]Probe UMI counts in boldface indicate positive values above background cutoffs. Abbreviations - HC: heavy chain; LC: light chain; UMI: unique molecular identifier.

**Table S2:** DENV-specificity of B cell subsets identified by LIBRA-seq.

| | DENV+ count | DENV- count | Freq. DENV+ |
|---|---|---|---|
| **DEN1** | **243** | **5297** | **4.4%** |
| Naive B cells | 175 | 4100 | 4.1% |
| Non-switched memory B cells | 47 | 617 | 7.1% |
| Switched memory B cells | 15 | 411 | 3.5% |
| Exhausted B cells | 6 | 169 | 3.4% |
| **DEN12** | **323** | **3661** | **8.1%** |
| Naive B cells | 38 | 911 | 4.0% |
| Non-switched memory B cells | 75 | 1043 | 6.7% |
| Switched memory B cells | 143 | 1148 | 11.1% |
| Exhausted B cells | 67 | 558 | 10.7% |
| **DEN24** | **154** | **6054** | **2.5%** |
| Naive B cells | 103 | 3548 | 2.8% |
| Non-switched memory B cells | 24 | 540 | 4.3% |
| Switched memory B cells | 21 | 1384 | 1.5% |
| Exhausted B cells | 6 | 580 | 1.0% |
| **Grand Total** | **720** | **15012** | **4.6%** |

**Table S3:** Reagents for E dimer probe competition cytometric bead assay.

| Marker | Fluorochrome | Clone | Catalog Number | Vendor | Final Dilution |
|---|---|---|---|---|---|
| Streptavidin (D1) | BUV563 | - | 612935 | BD Biosciences | 2ug/mL |
| Streptavidin (D2) | AF647 | - | S21374 | Invitrogen | 2ug/mL |
| Streptavidin (D3) | BV421 | - | 405225 | BioLegend | 2ug/mL |
| Streptavidin (D4) | RB-744 | - | 570516 | BD Biosciences | 2ug/mL |
| Streptavidin (ΔFL D2) | AF488 | - | S11223 | Invitrogen | 2ug/mL |
| **Antigens** | | | **Catalog Number** | **Vendor** | **Amount** |
| DENV1 E Dimer (I2-U6-P4-S1-Avi) | | | Custom | UNC Protein | 5ug/mL |
| DENV2 E Dimer (EV8 I2-U6-Avi) | | | Custom | UNC Protein | 5ug/mL |
| DENV3 E Dimer (I9-U6-P4-S1-Avi) | | | Custom | UNC Protein | 5ug/mL |
| DENV4 E Dimer (I2-U6-P4-S1-Avi) | | | Custom | UNC Protein | 5ug/mL |
| ΔFL DENV2 E Dimer (I2-I8-U6-Avi) | | | Custom | UNC Protein | 5ug/mL |
| D-Biotin | | | B20656 | Invitrogen | 200uM |

**Table S4:** Antibodies and reagents for immunophenotyping spectral flow panel.

| Marker | Fluorochrome | Clone | Catalog Number | Vendor | Final Dilution |
|---|---|---|---|---|---|
| CD8 | AF350 | RPA-T8 | FAB3802U | R&D Systems | 20 |
| CD45 | StarBright UV 510 | F10-89-4 | MCA87SBUV510 | Bio-Rad | 1250 |
| CD45RA | Spark UV 387 | HI100 | 304179 | BioLegend | 160 |
| CD69 | BUV563 | FN50 | 748764 | BD Biosciences | 80 |
| CD138 | BUV615 | MI15 | 751148 | BD Biosciences | 80 |
| CD27 | BUV661 | O323 | 751680 | BD Biosciences | 80 |
| IgG | BUV737 | G18-145 | 612819 | BD Biosciences | 13 |
| CD4 | BUV805 | SK3 | 612887 | BD Biosciences | 40 |
| Streptavidin (DENV3) | BV421 | - | 405225 | BioLegend | 2ug/mL |
| CD14 | BV510 | MphiP9 | 563079 | BD Biosciences | 160 |
| IgM | BV570 | MHM-88 | 314518 | BioLegend | 20 |
| CCR7 | BV605 | G043H7 | 314518 | BioLegend | 30 |
| CD71 | BV650 | CY1G4 | 334116 | BioLegend | 80 |
| CD184 (CXCR4) | BV711 | 12G5 | 740799 | BD Biosciences | 30 |
| CD185 (CXCR5) | BV750 | RF8B2 | 569501 | BD Biosciences | 60 |
| IgD | BV785 | IA6-2 | 348242 | BioLegend | 320 |
| IgA | AF488 | HISA43 | NBP3-11510AF488 | Novus Biologicals | 320 |
| HLA-DR | RB-545 | G46-6 | 756637 | BD Biosciences | 640 |
| CD38 | StarBright Blue 615 | AT13/5 | MCA1019SBB615 | Bio-Rad | 320 |
| CD16 | NFB660\120S | CB16 | H006T03B08-A | eBioscience | 80 |
| CD278 (PD-1) | RB-705 | EH12.1 | 570246 | BD Biosciences | 40 |
| Streptavidin (DENV4) | RB-744 | - | 570516 | BD Biosciences | 2ug/mL |
| CD183 (CXCR3) | RB-780 | 1C6 | 755404 | BD Biosciences | 60 |
| CD19 | RY-586 | SJ25C1 | 568108 | BD Biosciences | 1280 |
| Streptavidin (DENV1) | AF594 | - | 405240 | BioLegend | 2ug/mL |
| CD3 | RY703 | UCHT1 | 571431 | BD Biosciences | 40 |
| CD24 | NFY755 | SN3 | H064T02Y08-N | Invitrogen | 80 |
| CD32b | PE-Cy7 | S18005H | 398314 | BioLegend | 320 |
| Streptavidin (DENV2) | AF647 | - | S21374 | Invitrogen | 2ug/mL |
| Streptavidin (CoV2) | AF700 | - | S21383 | Invitrogen | 2ug/mL |
| Live/Dead | Zombie NIR | - | 423106 | BioLegend | 1600 |
| CD20 | APC-Fire 750 | 2H7 | 302358 | BioLegend | 320 |
| **Staining Buffers** | | | **Catalog Number** | **Vendor** | **Volume** |
| BD Horizon™ Brilliant Stain Buffer Plus | | | 566385 | BD Biosciences | 20uL |
| True-Stain Monocyte Blocker™ | | | 426101 | BioLegend | 5uL |
| Human TruStain FcX™ | | | 422302 | BioLegend | 5uL |
| CellBlox™ Plus Blocking Buffer | | | C001T06F01 | Invitrogen | 5uL |
| **Antigens** | | | **Catalog Number** | **Vendor** | **Amount** |
| DENV1 E Dimer (I2-U6-P4-S1-Avi) | | | Custom | UNC Protein | 5ug/mL |
| DENV2 E Dimer (EV8 I2-U6-Avi) | | | Custom | UNC Protein | 5ug/mL |
| DENV3 E Dimer (I9-U6-P4-S1-Avi) | | | Custom | UNC Protein | 5ug/mL |
| DENV4 E Dimer (I2-U6-P4-S1-Avi) | | | Custom | UNC Protein | 5ug/mL |
| Biotinylated HSA Protein, His, Avitag™ | | | HSA-H82E3-25ug | AcroBiosystems | 4ug/mL |
| Biotinylated SARS-CoV-2 S protein (D614G), His,Avitag™ | | | SPN-C82E3-25ug | AcroBiosystems | 4ug/mL |
| D-Biotin | | | B20656 | Invitrogen | 200uM |
